# Emulating randomized controlled trials of long-acting insulins and cardiovascular events using real-world data for patients with type 2 diabetes

**DOI:** 10.1101/2024.11.28.24317254

**Authors:** Wanning Wang, Pauline Reynier, Michael Webster-Clark, Oriana HY Yu, Vanessa Brunetti, Kristian B. Filion

**Affiliations:** Department of Epidemiology, Biostatistics and Occupational Health, McGill University, Montreal, Quebec, Canada; Center for Clinical Epidemiology, Lady Davis Institute, Jewish General Hospital, Montreal, Quebec, Canada; Department of Medicine, McGill University, Montreal, Quebec, Canada; Division of Endocrinology and Metabolism, Jewish General Hospital/McGill University, Montreal, Quebec, Canada; Center for Observational Research, Amgen Ltd, Uxbridge, UK

## Abstract

**Aims:** Randomized controlled trials (RCTs) have high internal validity but often have limited generalizability. To our knowledge, there are no studies examining potential differences between RCTs and real-world data in patient characteristics and risk of major adverse cardiovascular events (MACE) among patients with type 2 diabetes mellitus (T2DM) treated with long-acting insulin analogues.

**Methods:** We emulated the DEVOTE trial of insulin degludec vs glargine among patients with T2DM using data from the United Kingdom’s Clinical Practice Research Datalink. DEVOTE eligible and ineligible subpopulations were created. Cox proportional hazards models with inverse probability of treatment weighting were used to estimate hazard ratios (HRs) and corresponding confidence intervals (CIs) for MACE comparing new users of insulin degludec to new users of insulin glargine overall and in the eligible/ineligible subpopulations.

**Results:** There were 10,430 patients in the overall population, 5,280 in the DEVOTE eligible population, and 5,150 in the DEVOTE ineligible population. The overall (HR: 1.36, 95% CI: 0.83, 1.86) and DEVOTE eligible populations (HR: 1.07, 95% CI: 0.63, 1.58) were compatible with findings from the DEVOTE trial (HR: 0.91, 95% CI: 0.78, 1.06) for the risk of MACE. Due to a low number of events the DEVOTE ineligible population had deviations in point estimates and wider CIs (HR: 2.19, 95% CI: 0.30, 3.83).

**Conclusion:** The risk of MACE among patients with T2DM newly prescribed insulin degludec compared to insulin glargine was consistent between the overall population and the DEVOTE eligible subpopulation, while the DEVOTE ineligible population had discrepant point estimates.

**Twitter Summary:** Our study emulated the DEVOTE trial using RWD. Half of the RWD population would be eligible for the trial. RWD and RCTs had compatible effect estimate for risk of major cardiovascular outcomes.

## INTRODUCTION

Insulin is prescribed to patients with type 2 diabetes mellitus (T2DM) when other antidiabetic drugs have failed to achieve or maintain glycemic control and to prevent micro- (retinopathy, neuropathy and nephropathy) and macrovascular (cardiovascular) complications^1,2^. There are two main types of insulin, human insulin (often neutral protamine hagedorn) and long-acting insulin analogues (glargine, degludec, detemir)^3^. The efficacy of long-acting insulin analogues for the prevention of adverse cardiovascular events has been examined in two large randomized controlled trials (RCTs)^4,5^. However, the inclusion criteria of these trials were highly selective and may have resulted in trial populations that lack representativeness of characteristics of patients using long-acting insulin analogues in everyday clinical practice. For example, the ORIGIN trial selected individuals with pre-diabetes only^5^. In the DEVOTE trial^4^, which compared cardiovascular safety of insulin degludec and insulin glargine, the inclusion criteria were based on blood glucose levels and the presence of elevated cardiovascular risk factors; while this increased the number of events (and thus reduced the sample size required to achieve adequate statistical precision), it also makes the generalizability of its findings to patients without elevated cardiovascular risk unclear.

As real-world data (RWD) generated from electronic health records, claims data, and registries are becoming readily available, there has been a push by regulatory and health technology assessment agencies to utilize real-world evidence (RWE) in conjunction with RCTs to evaluate the effectiveness and safety of medical interventions^6–8^. Some studies examining the safety and efficacy of treatments of T2DM have produced concordant findings between RCTs and observational studies^9^. However, there is currently no literature comparing differences in patient characteristics and outcomes between RCTs and real-world populations among patients with T2DM using long-acting insulin analogues. The objective of this study was to assess the generalizability and representativeness of DEVOTE by examining the effect of long-acting insulin analogues on cardiovascular outcomes among patients with T2DM using RWD overall and in subpopulations defined by DEVOTE trial eligibility.

## METHODS

### Data Source

We conducted an observational study emulating a RCT using population-based data from the United Kingdom’s (UK) Clinical Practice Research Database (CPRD) Aurum. CPRD Aurum consists of primary care records of over 41 million patients in the UK, accounting for ∼20% of the population^10^. It contains longitudinal routinely collected electronic health records, with information on demographic characteristics, diagnoses and symptoms, drug exposures, laboratory tests, and referrals to hospital and specialist care. The CPRD is representative of the English population regarding geographical spread, socioeconomic deprivation, age, and gender^11^. Given the rich primary care data, CPRD Aurum is well suited for studies of people with T2DM, as T2DM is predominantly treated by general practitioners in the UK^12^. CPRD diagnoses and non-prescription information are coded using SNOMED CT (UK Edition), Read Version 2, and local EMIS Web ® codes. Drug and device prescriptions are coded using the Dictionary of Medicines and Devices and stored in the EMIS Web ® electronic medical record^11^. CPRD Aurum data were linked to Hospital Episode Statistics (HES) Admitted Patient Care (APC) and Office of National Statistics (ONS) death registration data. HES APC provides hospitalization data in the UK, including date of admission, date of discharge and diagnoses made during hospital stay^13^. ONS death registration data contain records of all UK deaths, with the deceased’s age, sex, and cause of death. HES APC and ONS diagnoses and causes of death are coded using the International Classification of Disease and Related Health Problems (ICD) 10^th^ revision.

The protocol of this study (19_217) was approved by the CPRD’s Independent Scientific Advisory Committee (ISAC) and by the CIUSSS West-Central Montreal Research Ethics Board (Montréal, Canada).

### Study population

The study population included patients diagnosed with T2DM who were prescribed insulin degludec or insulin glargine between March 1^st^, 2013 (when degludec became available in the UK) and November 31^st^, 2018 (end of data availability). The cohort was restricted to patients newly treated with insulin glargine or degludec, including those who switched from an oral antidiabetic medication or from another basal insulin to a treatment of interest. The date of the first prescription of insulin glargine or degludec defined study cohort entry. We excluded patients that had 1) < 1 year of recorded medical history (to capture comorbidities); 2) age <18 years; 3) previous diagnosis of type 1 diabetes (to ensure we capture only patients with T2DM); 4) diagnosis of gestational diabetes in the previous year (to ensure we captured only patients with T2DM); 5) zero days of follow-up (to capture outcomes); and 6) patients prescribed both long-acting insulin analogues on the day of cohort entry (to prevent mixing effects). Patients were followed until they experienced an outcome of interest (described below), censoring due to death of any cause, end of registration with CPRD, or the end of study period (November 31^st^, 2018), whichever occurred first. The population was limited to those with linkage between CPRD, HES, and ONS.

Using the cohort described above, we created two sub-populations; a DEVOTE eligible subpopulation and a DEVOTE ineligible subpopulation determined by the inclusion criteria of the DEVOTE trial. **Supplemental Table S1** outlines the operationalization of the RCT inclusion criteria for the CPRD and the number of patients that met each criterion.

### Exposure

In the DEVOTE trial, participants were randomized to insulin degludec or insulin glargine. We replicated this comparison, using glargine as an active comparator. In addition, we used an intention-to-treat (ITT) approach for exposure definition to reflect the study design and analysis used in the DEVOTE trial. An ITT approach is the suggested framework for studies that emulate RCT with observational data to ensure a useful treatment effect to compare to RCTs^14,15^. To mimic the ITT analyses of a trial, we used a time-fixed exposure definition where patients were classified into one of two mutually exclusive categories (new user of insulin degludec or new user of insulin glargine) from study entry until the end of follow-up regardless of treatment switching or discontinuation. For all exposure categories, use of other antidiabetic drugs including other insulins were permitted.

### Outcome

Our primary outcome was the occurrence of major adverse cardiovascular events (MACE), a composite endpoint of myocardial infarction (MI), ischemic stroke, or cardiovascular death. For our secondary outcomes, we examined the individual components of MACE, all-cause mortality, hospitalization for heart failure (HHF), and hospitalization for hypoglycemia. HHF and hypoglycemia were not reported in the DEVOTE trial but were included as they were important safety signals from previous CVOTs of other antidiabetic medications^16^. We used the following ICD-10 codes in HES and ONS to define the outcomes of interest: MI (ICD-10: I21.x), ischemic stroke (ICD-10: I63.x-I64.x), and HHF (ICD-10: I11.0, I13.0, I13.2, I50.x). The event date was defined by the date of admission for HES-defined events and date of death for ONS-defined events. Death from cardiovascular disease was defined using the underlying cause of death in ONS (ICD-10: I00.x-I78.x [except 146.9]). All-cause mortality was defined using CPRD, HES, and ONS, with the earliest recorded date of death defining the event date. Hypoglycemia was defined by a relevant ICD-10 code in HES (E16.0, E16.1, E16.2).

### Covariates

We included three types of baseline covariates in our propensity score (PS) model: demographics, comorbidities, and medications used at cohort entry. Covariates measured at baseline include year of cohort entry, age, sex, race, Index of Multiple Deprivation deciles, duration of treated diabetes (time since first prescription for an antidiabetic drug and cohort entry date), smoking status, previous history of alcohol related disorders, atrial fibrillation, previous diagnosis of cancer (not including non-melanoma skin cancer), chronic obstructive pulmonary disease, acute kidney injury, chronic kidney disease, retinopathy, neuropathy, dialysis, cerebrovascular disease, body mass index (BMI) (last measurement before cohort entry), A1C (last measured before cohort entry), and estimated glomerular filtration rate (eGFR). The medications assessed at baseline were ACE inhibitors, angiotensin II receptor blockers, beta blockers, diuretics, statins, acetylsalicylic acid, non-steroidal anti-inflammatory drugs, fibrates, and use of other diabetic medications (metformin, sulfonylureas, thiazolidinediones, DPP-4 inhibitors, GLP-1 receptor agonists, alpha-glucosidase inhibitors, meglitinides, SGLT-2 inhibitors, and other types of insulin). For our MACE models, we further adjusted for coronary artery disease, hyperlipidemia, hypertension, peripheral vascular disease, stroke, MI, coronary revascularization, systolic blood pressure (latest measure before cohort entry), diastolic blood pressure (latest measure before cohort entry), total cholesterol, use of calcium channel blockers, oral anticoagulants, and antiplatelets. For the hypoglycemia models, we also included history of hospitalization for hypoglycemia in the year prior to cohort entry, thyroid disease (including hypothyroidism and hyperthyroidism), liver cirrhosis, previous medication usages of acetaminophen, opioids, and glucagon.

We used multiple imputation by chained equations (MICE) to impute missing values for the variables of race, Index of Multiple Deprivation decile, smoking status, A1C, eGFR, BMI, and systolic and diastolic blood pressure. Missingness for these variables was less than 5%.

### Statistical Analyses

We compared the patient characteristics between those eligible and ineligible for the DEVOTE trial and our overall population to the DEVOTE trial using absolute values of the standardized difference. Standardized differences greater than 0.1 were considered important^17^. We used proportions with 95% confidence intervals (CIs) to calculate the real-world populations who would have been eligible for the trial.

We used PS to account for differential baseline covariates between study groups. All covariates mentioned above were included in our model using multivariable logistic regression, with the inverse of the computed PS subsequently used to weigh exposure groups via inverse probability of treatment weights (IPTW). We estimated absolute values of standardized differences to compare the characteristics of each exposure group before and after weighting. After stabilized IPTW, we truncated weights at 10 to minimize the impact of extreme weights^18^. We used Poisson regression to calculate crude incidence rates and 95% CIs for each subpopulation for the primary and secondary outcomes. Using the exposure groups created after IPTW, time-fixed Cox proportional hazards models were used to estimate hazard ratios (HRs) and bootstrapped 95% CIs (by finding the 2.5th and 97.5th percentiles) for MACE in our three study populations.

We performed two secondary analyses. First, we repeated our analysis for individual components of MACE as well as all-cause mortality, HHF, and hospitalization due to hypoglycemia. Second, similar to the DUPLICATE studies^9,19^, we estimated three agreement statistics (statistical agreement, estimate agreement and standardized difference agreement) to compare our RWD to RCTs for all three of our populations. The definitions for these agreement statistics can be found in the **Supplemental Methods.** We conducted six sensitivity analyses to examine the robustness of our results; these analyses are described in the **Supplemental Methods.**

### RESULTS

**Figure 1** describes our study cohort composition. Our CPRD cohort consisted of 10,430 patients with T2DM, of which 812 initiated insulin degludec and 9,618 initiated insulin glargine. From our CPRD cohort, 5,280 patients were eligible for the DEVOTE trial (51% [95% CI: 49%, 52%] of the total population). The mean follow-up time was 1.5 years among insulin degludec initiators and 2.1 years among insulin glargine initiators. **Table 1** and **Supplemental Table S2** describe baseline patient characteristics for each exposure group. Across all subpopulations, degludec users were younger, had higher BMI, and greater use of other anti-diabetic medications. **Table 2** and **Supplemental Table S3** describe absolute standardized mean differences between populations after multiple imputation and weighting. After imputation and weighting, most characteristics had a standardized difference of <0.1. **Supplemental Figure S2** and **Supplemental Figure S3** illustrate the absolute standardized mean differences between the DEVOTE eligible and DEVOTE ineligible subpopulations and between the trial population and the RWD populations, respectively, demonstrating large degrees of heterogeneity in patient characteristics.

**Figure 1.**
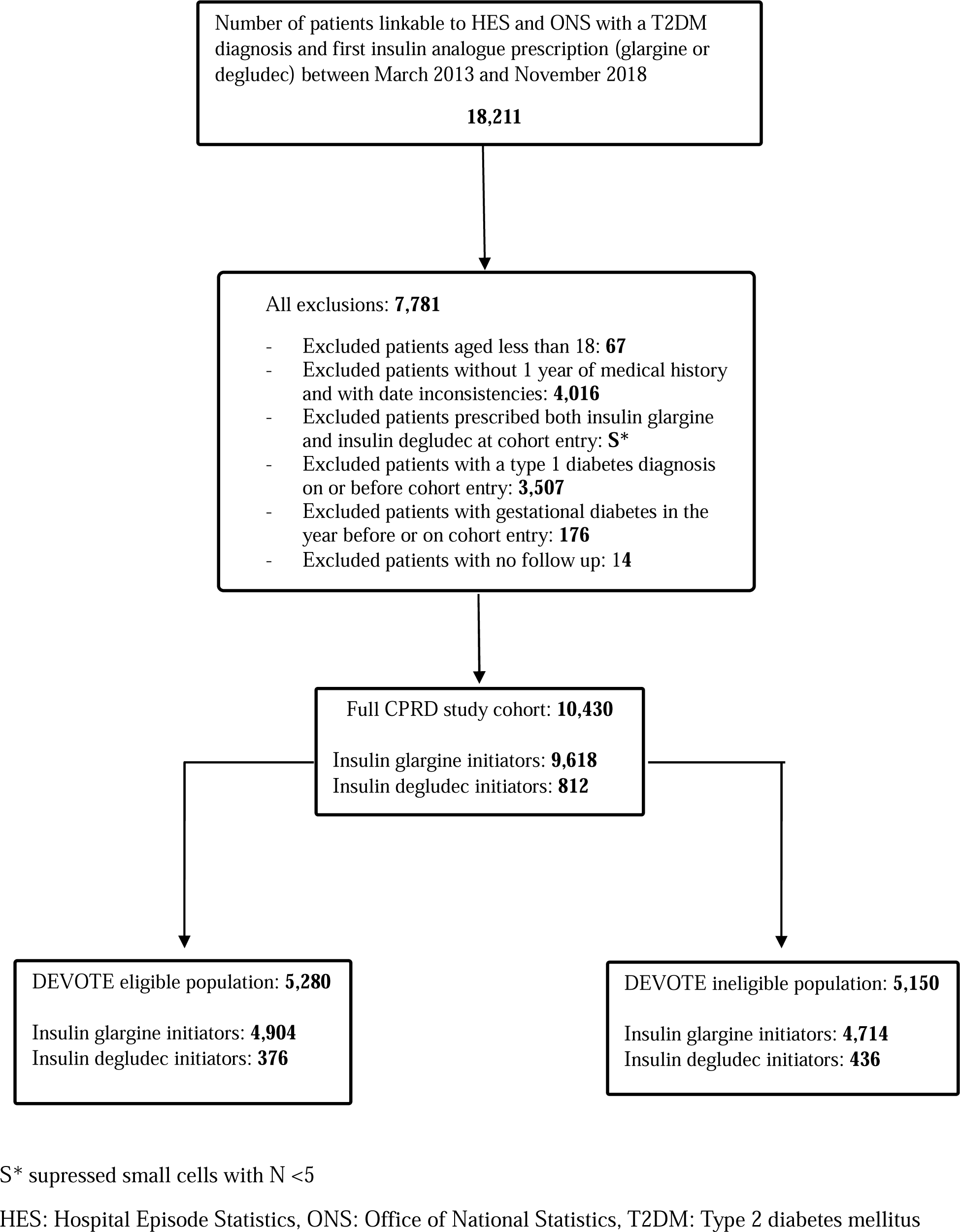
Flowchart of cohort to define sub-populations and exposures group of insulin glargine and insulin degludec initiators to examine risk of MACE in patients with type 2 diabetes.

**Table 1.** Baseline characteristics of CPRD, DEVOTE eligible and DEVOTE ineligible populations of patients with type 2 diabetes who initiated insulin degludec or insulin glargine before inverse probability of treatment weighting and multiple imputation.

|  | CPRD |  |  | DEVOTE eligible |  |  | DEVOTE ineligible |  |  |
| --- | --- | --- | --- | --- | --- | --- | --- | --- | --- |
|  | Insulin<br>glargine<br>users<br>N/mean<br>(%/SD) | Insulin<br>degludec<br>users<br>N/mean<br>(%/SD) | SMD | Insulin<br>glargine<br>users<br>N/mean<br>(%/SD) | Insulin<br>degludec<br>users<br>N/mean<br>(%/SD) | SMD | Insulin<br>glargine<br>users<br>N/mean<br>(%/SD) | Insulin<br>degludec<br>users<br>N/mean<br>(%/SD) | SMD |
| Number of patients | 9,618 | 812 |  | 4,904 | 376 |  | 4,714 | 436 |  |
| Age (years) Mean (SD) | 63.9 (14.1) | 60.5 (12.9) | 0.25 | 71.8 (10.3) | 68.1 (10.0) | 0.38 | 55.6 (12.9) | 54.1 (11.6) | 0.13 |
| Males, n (%) | 5,381 (55.9) | 443 (54.6) | 0.03 | 2,724 (55.5) | 210 (55.9) | 0.01 | 2,657 (56.4) | 233 (53.4) | 0.06 |
| Ethnicity, n (%) |  |  |  |  |  |  |  |  |  |
| Caucasian | 7,440 (77.4) | 664 (81.8) | 0.19 | 4,103 (83.7) | 332 (88.3) | 0.18 | 3,337 (70.8) | 332 (76.1) | 0.21 |
| Non-Caucasian | 1,403 (14.6) | 70 (8.6) |  | 628 (12.8) | 28 (7.4) |  | 775 (16.4) | 42 (9.6) |  |
| Year of Cohort Entry, n (%) |  |  |  |  |  |  |  |  |  |
| 2013 | 1,024 (10.6) | 19 (2.3) | 0.34 | 532 (10.6) | 10 (2.7) | 0.33 | 492 (10.4) | 9 (2.1) | 0.35 |
| 2014 | 1,298 (13.5) | 32 (3.9) | 0.34 | 658 (13.4) | 12 (3.2) | 0.38 | 640 (13.6) | 20 (4.6) | 0.32 |
| 2015 | 1,561 (16.2) | 78 (9.6) | 0.20 | 749 (15.3) | 37 (9.8) | 0.16 | 812 (17.2) | 41 (9.4) | 0.24 |
| 2016 | 1,783 (18.5) | 149 (18.3) | 0.01 | 907 (18.5) | 74 (19.7) | 0.03 | 876 (18.6) | 75 (17.2) | 0.04 |
| 2017 | 2,008 (20.9) | 256 (31.5) | 0.23 | 1,029 (21.0) | 120 (31.9) | 0.25 | 979 (20.8) | 136 (31.2) | 0.24 |
| 2018 | 1,944 (20.2) | 278 (7.6) | 0.32 | 1,029 (21.0) | 123 (32.7) | 0.27 | 915 (19.4) | 155 (35.6) | 0.37 |
| Diabetes duration (years) |  |  |  |  |  |  |  |  |  |
| Mean (SD)* | 10.8 (7.0) | 11.6 (6.1) | 0.12 | 13.2 (6.9) | 13.6 (6.1) | 0.06 | 8.4 (6.2) | 9.9 (5.5) | 0.25 |
| Lifestyle and other covariates |  |  |  |  |  |  |  |  |  |
| BMI, Mean (SD)<sup>†</sup> | 31.0 (7.0) | 33.5 (7.5) | 0.35 | 30.8 (6.7) | 33.3 (7.1) | 0.37 | 31.2 (7.3) | 33.6 (7.8) | 0.32 |
| Smoking, n (%)<sup>†</sup> |  |  |  |  |  |  |  |  |  |
| Current | 3,588 (37.3) | 292 (36.0) | 0.03 | 1,830 (37.3) | 128 (34.0) | 0.07 | 1,758 (37.3) | 164 (37.8) | 0.00 |
| Former | 3,174 (33.0) | 300 (36.9) | 0.08 | 1,735 (35.4) | 151 (40.2) | 0.10 | 1,439 (30.5) | 149 (34.2) | 0.07 |
| Never | 2,699 (28.1) | 213 (26.2) | 0.05 | 1,282 (26.1) | 94 (25.0) | 0.03 | 1,417 (30.1) | 119 (27.2) | 0.07 |
| Systolic Blood Pressure Level, | 131.1 (15.8) | 131.1 (14.2) | 0.00 | 131.7 (16.4) | 131.0 (14.9) | 0.04 | 130.5 (15.1) | 131.2 (13.5) | 0.04 |
| Mean (SD)<sup>†</sup> |  |  |  |  |  |  |  |  |  |
| Diastolic Blood Pressure Level, Mean (SD)<sup>†</sup> | 75.2 (10.0) | 75.4 (9.4) | 0.02 | 73.1 (9.9) | 73.2 (9.5) | 0.01 | 77.5 (9.5) | 77.3 (8.9) | 0.02 |
| A1C, Mean (SD)<sup>†</sup> | 9.8 (2.2) | 9.9 (2.0) | 0.01 | 9.7 (2.2) | 9.9 (2.0) | 0.08 | 10.0 (2.2) | 9.9 (2.0) | 0.06 |
| eGFR, Mean (SD)<sup>†</sup> | 66.5 (21.3) | 70.9 (19.0) | 0.22 | 57.2 (21.6) | 62.5 (19.9) | 0.26 | 76.6 (15.7) | 78.2 (14.7) | 0.11 |
| Comorbidities, n (%) |  |  |  |  |  |  |  |  |  |
| Acute kidney injury | 1,612 (16.8) | 93 (11.5) | 0.15 | 1,263 (25.8) | 79 (21.0) | 0.11 | 349 (7.4) | 14 (3.2) | 0.19 |
| Alcohol related disorders | 1,937 (20.1) | 140 (17.2) | 0.07 | 988 (20.1) | 63 (16.8) | 0.09 | 949 (20.1) | 77 (17.7) | 0.06 |
| Atrial fibrillation | 782 (8.1) | 59 (7.3) | 0.03 | 687 (14.0) | 50 (13.3) | 0.02 | 95 (2.0) | 9 (2.1) | 0.00 |
| Cancer<sup>‡</sup> | 2,340 (24.3) | 147 (18.1) | 0.15 | 1,444 (29.4) | 91 (24.2) | 0.12 | 896 (19.0) | 56 (12.8) | 0.17 |
| Cerebrovascular disease | 1,303 (13.5) | 79 (9.7) | 0.12 | 1,146 (23.4) | 72 (19.1) | 0.10 | 157 (3.3) | 7 (1.6) | 0.11 |
| Chronic kidney disease | 2,683 (27.9) | 155 (19.1) | 0.21 | 2,480 (50.6) | 141 (37.5) | 0.27 | 203 (4.3) | 14 (3.2) | 0.06 |
| COPD | 1,102 (11.5) | 72 (8.9) | 0.09 | 811 (16.5) | 54 (14.4) | 0.06 | 291 (6.2) | 18 (4.1) | 0.09 |
| Coronary artery disease | 2,853 (29.7) | 216 (26.6) | 0.07 | 2,606 (53.1) | 204 (54.3) | 0.02 | 247 (5.2) | 12 (2.8) | 0.13 |
| Coronary revascularization | 781 (8.1) | 55 (6.8) | 0.05 | 716 (14.6) | 53 (14.1) | 0.01 | 65 (1.4) | S* | - |
| Dialysis | 152 (1.6) | 15 (1.8) | 0.02 | 96 (2.0) | 12 (3.2) | 0.08 | 56 (1.2) | S* | - |
| Hyperlipidemia | 4,184 (43.5) | 347 (42.7) | 0.02 | 2,757 (56.2) | 214 (56.9) | 0.01 | 1,427 (30.3) | 133 (30.5) | 0.01 |
| Hypertension | 7,069 (73.5) | 607 (74.8) | 0.03 | 4,370 (89.1) | 342 (91.0) | 0.06 | 2,699 (57.3) | 265 (60.8) | 0.07 |
| Hypoglycemia | 1,011 (10.5) | 89 (11.0) | 0.01 | 624 (12.7) | 51 (13.6) | 0.02 | 387 (8.2) | 38 (8.7) | 0.02 |
| Myocardial Infarction | 2,228 (23.2) | 165 (20.3) | 0.07 | 2,031 (41.4) | 157 (41.8) | 0.01 | 197 (4.2) | 8 (1.8) | 0.14 |
| Neuropathy | 2,588 (26.9) | 206 (25.4) | 0.04 | 1,585 (32.3) | 112 (29.8) | 0.05 | 1,003 (21.3) | 94 (21.6) | 0.01 |
| Peripheral vascular disease | 1,174 (12.2) | 92 (11.3) | 0.03 | 1,020 (20.8) | 82 (21.8) | 0.02 | 154 (3.3) | 10 (2.3) | 0.06 |
| Retinopathy | 4,454 (46.3) | 409 (50.4) | 0.08 | 2,676 (54.6) | 224 (59.6) | 0.10 | 1,778 (37.7) | 185 (42.4) | 0.10 |
| Stroke | 988 (10.3) | 71 (8.7) | 0.05 | 896 (18.3) | 70 (18.6) | 0.01 | 92 (2.0) | S* | - |
| Cirrhosis | 168 (1.7) | 7 (0.9) | 0.08 | 90 (1.8) | 6 (1.6) | 0.02 | 78 (1.7) | S* | - |
| Antidiabetic medications, n (%) |  |  |  |  |  |  |  |  |  |
| Metformin | 7,427 (77.2) | 646 (79.6) | 0.06 | 3,617 (73.8) | 279 (74.2) | 0.01 | 3,810 (80.8) | 367 (84.2) | 0.09 |
| Sulfonylureas | 5,889 (61.2) | 388 (47.8) | 0.27 | 3,199 (65.2) | 185 (49.2) | 0.33 | 2,690 (57.1) | 203 (46.6) | 0.21 |
| Thiazolidinediones | 626 (6.5) | 45 (5.5) | 0.04 | 338 (6.9) | 18 (4.8) | 0.09 | 288 (6.1) | 27 (6.2) | 0.00 |
| DPP-4 inhibitors | 3,996 (41.5) | 259 (31.9) | 0.20 | 2,291 (46.7) | 127 (33.8) | 0.27 | 1,705 (36.2) | 132 (30.3) | 0.13 |
| GLP-1 receptor agonists | 1,571 (16.3) | 414 (51.0) | 0.79 | 653 (13.3) | 169 (44.9) | 0.74 | 918 (19.5) | 245 (56.2) | 0.82 |
| Alpha-glucosidase inhibitors | 37 (0.4) | S* | - | 23 (0.5) | S* | - | 14 (0.3) | S* | - |
| Meglitinides | 102 (1.1) | 7 (0.9) | 0.02 | 54 (1.1) | 6 (1.6) | 0.04 | 48 (1.0) | S* | - |
| SGLT-2 inhibitors | 1,212 (12.6) | 217 (26.7) | 0.36 | 438 (8.9) | 83 (22.1) | 0.37 | 774 (16.4) | 134 (30.7) | 0.34 |
| Insulin (bolus or long-acting not including insulin glargine and degludec) | 3,451 (35.9) | 429 (52.8) | 0.35 | 1,668 (34.0) | 219 (58.2) | 0.50 | 1,783 (37.8) | 210 (48.2) | 0.21 |

\*Duration of type 2 diabetes, defined as time since first diagnostic A1C, diagnostic code, or initiation of antihyperglycemic medication.
† Percentage of missing data presented in supplementary material, along with categorical presentation of the data.
‡Not including non-melanoma skin cancer
S\* suppressed small cells with N < 5
Abbreviations: A1C: glycated hemoglobin, BMI: body mass index, COPD: chronic obstructive pulmonary disorder, CPRD: Clinical Practice Research Datalink, DPP-4 inhibitors: dipeptidyl peptidase-4 inhibitors, eGFR: estimated glomerular filtration rate, GLP-1 receptor agonists: glucagon-like peptide-1 receptor agonists, SD: standard deviation, SGLT-2 inhibitors: sodium-glucose cotransporter-2 inhibitors, SMD: standardized mean difference

**Table 2.** Baseline characteristics of CPRD, DEVOTE eligible and DEVOTE ineligible populations of patients with type 2 diabetes who initiated insulin degludec or insulin glargine after inverse probability of treatment weighting and multiple imputation.

|  | CPRD |  |  | DEVOTE eligible |  |  | DEVOTE ineligible |  |  |
| --- | --- | --- | --- | --- | --- | --- | --- | --- | --- |
|  | Insulin<br>glargine<br>users | Insulin<br>degludec<br>users | SMD | Insulin<br>glargine<br>users | Insulin<br>degludec<br>users | SMD | Insulin<br>glargine<br>users | Insulin<br>degludec<br>users | SMD |
|  | N/mean<br>(%/SD) | N/mean<br>(%/SD) |  | N/mean<br>(%/SD) | N/mean<br>(%/SD) |  | N/mean<br>(%/SD) | N/mean<br>(%/SD) |  |
| Number of patients | 9,637 | 769 |  | 4,916 | 320 |  | 4,722 | 414 |  |
| Age (years) Mean (SD) | 63.6 (14.1) | 63.2 (12.7) | 0.03 | 71.5 (10.2) | 70.6 (9.4) | 0.09 | 55.5 (12.8) | 56.4 (12.2) | 0.03 |
| Males, n (%) | 5,380 (55.8) | 453 (58.9) | 0.06 | 2,732 (55.6) | 183 (57.0) | 0.03 | 2,650 (56.1) | 247 (59.6) | 0.07 |
| Ethnicity, n (%) |  |  |  |  |  |  |  |  |  |
| Caucasian | 8,139 (84.5) | 665 (86.4) | 0.06 | 4,287 (87.2) | 294 (91.7) | 0.15 | 3,851 (81.6) | 346 (83.5) | 0.05 |
| Non-Caucasian | 1,498 (15.5) | 104 (13.6) |  | 629 (12.8) | 27 (8.3) |  | 871 (18.4) | 68 (16.5) |  |
| Year of Cohort Entry, n (%) |  |  |  |  |  |  |  |  |  |
| 2013 | 962 (10.0) | 112 (14.6) | 0.14 | 504 (10.2) | 35 (11.1) | 0.03 | 459 (9.7) | 61 (14.6) | 0.15 |
| 2014 | 1,227 (12.7) | 103 (13.4) | 0.02 | 622 (12.7) | 33 (10.2) | 0.08 | 604 (12.8) | 61 (14.8) | 0.06 |
| 2015 | 1,509 (15.7) | 83 (10.8) | 0.14 | 729 (14.8) | 34 (10.5) | 0.13 | 779 (16.5) | 40 (9.6) | 0.20 |
| 2016 | 1,780 (18.5) | 126 (16.4) | 0.05 | 913 (18.6) | 65 (20.2) | 0.04 | 868 (18.4) | 57 (13.8) | 0.12 |
| 2017 | 2,090 (21.7) | 172 (22.3) | 0.01 | 1,069 (21.7) | 75 (23.4) | 0.04 | 1,022 (21.6) | 97 (23.4) | 0.04 |
| 2018 | 2,069 (21.5) | 173 (22.5) | 0.03 | 1,079 (22.0) | 79 (24.6) | 0.06 | 989 (21.0) | 98 (23.7) | 0.07 |
| Diabetes duration (years) |  |  |  |  |  |  |  |  |  |
| Mean (SD)* | 10.9 (7.0) | 10.8 (6.0) | 0.01 | 13.2 (6.9) | 13.5 (6.2) | 0.05 | 8.5 (6.2) | 9.1 (5.5) | 0.09 |
| Lifestyle and other covariates |  |  |  |  |  |  |  |  |  |
| BMI, Mean (SD)<sup>†</sup> | 31.2 (7.2) | 31.6 (6.7) | 0.07 | 31.0 (6.8) | 31.8 (5.9) | 0.14 | 31.3 (7.3) | 31.4 (7.0) | 0.01 |
| Smoking<sup>†</sup>, n (%) |  |  |  |  |  |  |  |  |  |
| Current | 3,640 (37.8) | 290 (37.6) | 0.00 | 1,843 (37.5) | 121 (37.9) | 0.01 | 1,801 (38.1) | 152 (36.8) | 0.03 |
| Former | 3,249 (33.7) | 290 (37.6) | 0.00 | 1,767 (35.9) | 111 (34.6) | 0.03 | 1,478 (31.3) | 130 (31.3) | 0.00 |
| Never | 2,748 (28.5) | 254 (33.1) | 0.01 | 1,306 (26.6) | 88 (27.5) | 0.02 | 1,444 (30.6) | 132 (31.9) | 0.03 |
| Systolic Blood Pressure Level, Mean (SD)<sup>†</sup> | 131.1 (15.8) | 132.8 (14.1) | 0.11 | 131.7 (16.3) | 132.1 (14.1) | 0.03 | 130.6 (15.1) | 132.6 (13.6) | 0.14 |
| Diastolic Blood Pressure Level, Mean (SD)<sup>†</sup> | 75.2 (9.9) | 75.7 (9.4) | 0.05 | 73.1 (9.9) | 72.7 (9.0) | 0.04 | 77.5 (9.5) | 78.0 (9.4) | 0.06 |
| A1C, Mean (SD)<sup>†</sup> | 9.9 (2.2) | 10.0 (2.0) | 0.07 | 9.7 (2.2) | 9.8 (1.9) | 0.02 | 10.0 (2.2) | 10.2 (2.0) | 0.10 |
| eGFR, Mean (SD)<sup>†</sup> | 67.0 (21.1) | 67.2 (19.6) | 0.01 | 57.7 (21.6) | 58.8 (19.0) | 0.05 | 76.7 (15.6) | 76.6 (15.3) | 0.01 |
| Comorbidities, n (%) |  |  |  |  |  |  |  |  |  |
| Acute kidney injury | 1,573 (16.3) | 124 (16.2) | 0.00 | 1,248 (25.4) | 85 (26.6) | 0.03 | 333 (7.0) | 32 (7.8) | 0.03 |
| Alcohol related disorders | 1,915 (19.9) | 164 (21.3) | 0.04 | 976 (19.8) | 66 (20.6) | 0.02 | 939 (19.9) | 92 (22.1) | 0.05 |
| Atrial fibrillation | 776 (8.1) | 58 (7.5) | 0.02 | 685 (13.9) | 44 (13.6) | 0.01 | 95 (2.0) | 8 (2.0) | 0.00 |
| Cancer<sup>‡</sup> | 2,299 (23.9) | 206 (26.7) | 0.07 | 1,428 (29.0) | 105 (32.6) | 0.08 | 875 (18.5) | 86 (20.7) | 0.05 |
| Cerebrovascular disease | 1,276 (13.2) | 112 (14.6) | 0.04 | 1,134 (23.1) | 86 (26.9) | 0.09 | 150 (3.2) | 7 (1.6) | 0.10 |
| Chronic kidney disease | 2,619 (27.2) | 185 (24.0) | 0.07 | 2,437 (49.6) | 150 (46.8) | 0.05 | 200 (4.2) | 13 (3.1) | 0.06 |
| COPD | 1,085 (11.3) | 95 (12.4) | 0.03 | 806 (16.4) | 54 (16.8) | 0.01 | 283 (6.0) | 24 (5.7) | 0.01 |
| Coronary artery disease | 2,836 (29.4) | 222 (28.9) | 0.01 | 2,616 (53.2) | 178 (55.6) | 0.05 | 237 (5.0) | 13 (3.2) | 0.09 |
| Coronary revascularization | 770 (8.0) | 51 (6.6) | 0.05 | 713 (14.5) | 43 (13.5) | 0.03 | 61 (1.3) | S* | - |
| Dialysis | 154 (1.6) | 11 (1.5) | 0.01 | 101 (2.0) | 6 (1.7) | 0.02 | 54 (1.1) | S* | - |
| Hyperlipidemia | 4,189 (43.5) | 321 (41.7) | 0.04 | 2,765 (56.3) | 182 (56.8) | 0.01 | 1,434 (30.4) | 130 (31.4) | 0.02 |
| Hypertension | 7,092 (73.6) | 545 (70.8) | 0.06 | 4,388 (89.3) | 292 (91.0) | 0.06 | 2,718 (57.6) | 221 (53.4) | 0.08 |
| Hypoglycemia | 1,019 (10.6) | 82 (10.6) | 0.00 | 626 (12.7) | 42 (13.2) | 0.02 | 396 (8.4) | 42 (10.1) | 0.06 |
| Myocardial Infarction | 2,210 (22.9) | 169 (22.0) | 0.02 | 2,036 (41.4) | 134 (41.9) | 0.01 | 187 (4.0) | 7 (1.7) | 0.14 |
| Neuropathy | 2,583 (26.8) | 234 (30.4) | 0.08 | 1,579 (32.1) | 105 (32.7) | 0.01 | 1,011 (21.4) | 107 (25.9) | 0.11 |
| Peripheral vascular disease | 1,167 (12.1) | 107 (13.9) | 0.05 | 1,023 (20.8) | 75 (23.4) | 0.06 | 151 (3.2) | 24 (5.8) | 0.13 |
| Retinopathy | 4,499 (46.7) | 356 (46.3) | 0.01 | 2,704 (55.0) | 197 (61.3) | 0.13 | 1,805 (38.2) | 142 (34.2) | 0.08 |
| Stroke | 979 (10.2) | 100 (13.1) | 0.09 | 901 (18.3) | 83 (25.8) | 0.18 | 85 (1.8) | S* | - |
| Cirrhosis | 162 (1.7) | 29 (3.8) | 0.13 | 90 (1.8) | 16 (5.1) | 0.18 | 72 (1.5) | S* | - |
| Antidiabetic medications, n (%) |  |  |  |  |  |  |  |  |  |
| Metformin | 7,461 (77.4) | 600 (78.0) | 0.01 | 3,627 (73.8) | 233 (72.8) | 0.02 | 3,833 (81.2) | 349 (84.2) | 0.08 |
| Sulfonylureas | 5,786 (60.0) | 435 (56.6) | 0.07 | 3,141 (63.9) | 175 (54.6) | 0.19 | 2,645 (56.0) | 236 (57.0) | 0.02 |
| Thiazolidinediones | 620 (6.4) | 45 (5.8) | 0.03 | 330 (6.7) | 15 (4.6) | 0.09 | 289 (6.1) | 26 (6.4) | 0.01 |
| DPP-4 inhibitors | 3,920 (40.7) | 296 (38.5) | 0.04 | 2,243 (45.6) | 124 (38.7) | 0.14 | 1,680 (35.6) | 152 (36.7) | 0.02 |
| GLP-1 receptor agonists | 1,851 (19.2) | 149 (19.4) | 0.00 | 775 (15.8) | 55 (17.3) | 0.04 | 1,074 (22.7) | 94 (22.8) | 0.00 |
| Alpha-glucosidase inhibitors | 35 (0.4) | S* | - | 22 (0.5) | S* | - | 13 (0.3) | S* | - |
| Meglitinides | 100 (1.0) | S* | - | 55 (1.1) | S* | - | 45 (0.9) | S* | - |
| SGLT-2 inhibitors | 1,332 (13.8) | 116 (15.0) | 0.03 | 492 (10.0) | 35 (11.0) | 0.03 | 838 (17.8) | 82 (19.8) | 0.05 |
| Insulin | 3,609 (37.5) | 377 (49.0) | 0.24 | 1,771 (36.0) | 174 (54.4) | 0.38 | 1,840 (39.0) | 173 (41.7) | 0.06 |

\* Duration of type 2 diabetes, defined as time since first diagnostic A1C, diagnostic code, or initiation of antihyperglycemic medication.
† Percentage of missing data presented in supplementary material, along with categorical presentation of the data.
‡ Not including non-melanoma skin cancer
S\* suppressed small cells with N < 5
Abbreviations: A1C: glycated hemoglobin, BMI: body mass index, COPD: chronic obstructive pulmonary disorder, CPRD: Clinical Practice Research Datalink, DPP-4 inhibitors: dipeptidyl peptidase-4 inhibitors, eGFR: estimated glomerular filtration rate, GLP-1 receptor agonists: glucagon-like peptide-1 receptor agonists, SD: standard deviation, SGLT-2 inhibitors: sodium-glucose cotransporter-2 inhibitors, SMD: standardized mean difference

The results of our primary analyses are reported in **Table 3**. In the overall CPRD population, the incidence rate of MACE was 39.4 events per 1000 person-years (95% CI: 29.6, 52.4) in the insulin degludec group and 45.5 events per 1000 person-years (95% CI: 42.6, 48.6) in the insulin glargine group. The incident rates for the DEVOTE eligible population were 71.4 events per 1000 person-years (95% CI: 52.2, 97.8) and 79.7 events per 1000 person-years (95% CI: 74.2, 85.6), respectively. The DEVOTE ineligible patients had lower incidence rates for both exposure groups (insulin degludec: 12.3 events per 1000 person-years, 95% CI: 6.2, 24.7; insulin glargine: 15.3 events per 1000 person-years, 95% CI: 13.1, 17.8). In the overall CPRD population, the adjusted HR for MACE with degludec versus glargine was 1.36 (95% CI 0.83, 1.86). The adjusted HR was 1.07 (95% CI: 0.63, 1.58) for the DEVOTE eligible population and 2.19 (95% CI: 0.30, 3.83) for DEVOTE ineligible population. The DEVOTE trial had a HR of 0.91 (95% CI: 0.78, 1.06).

**Table 3.**
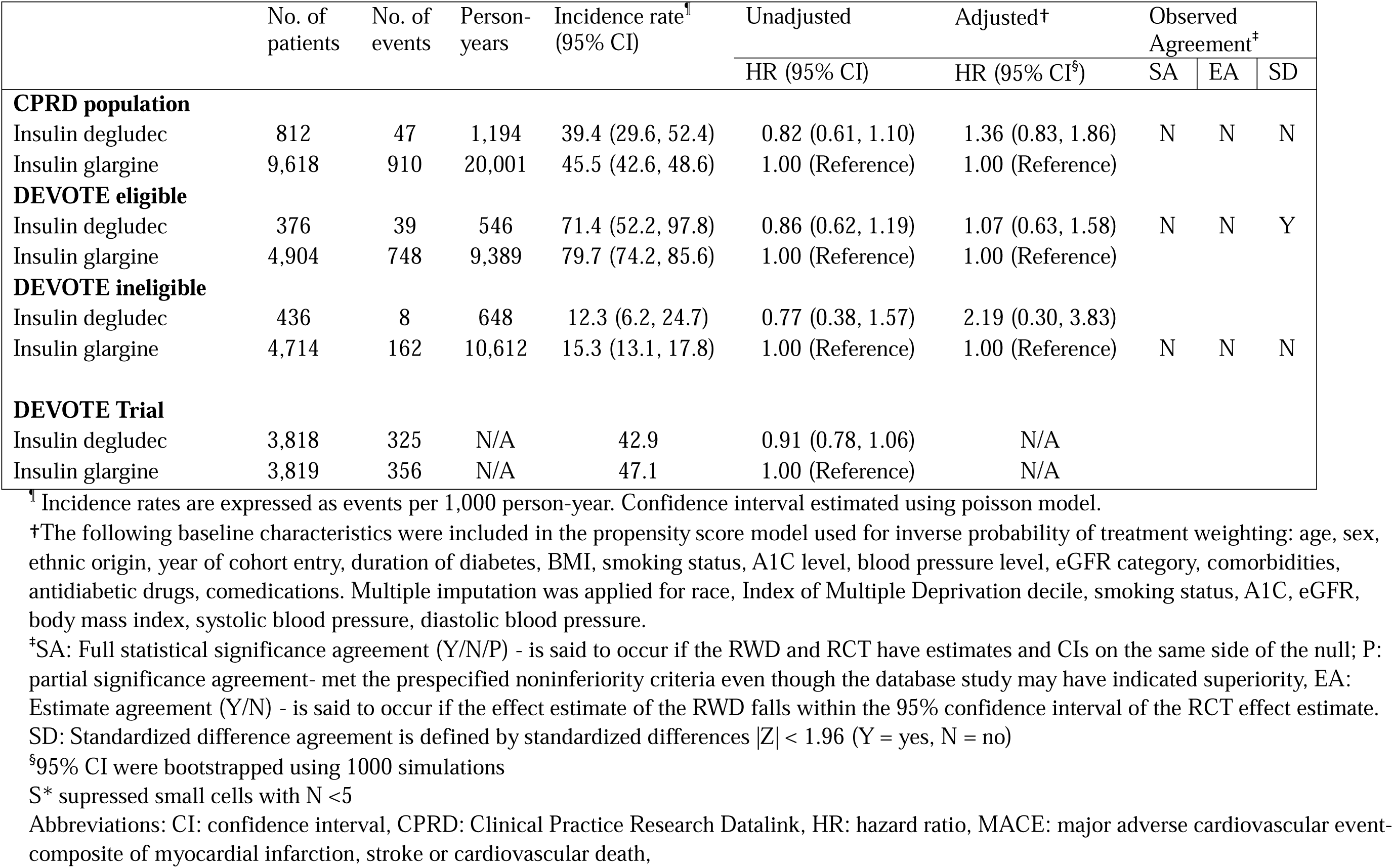
The association between use of insulin degludec compared to insulin glargine and the risk of MACE among patients with type 2 diabetes in the CPRD, DEVOTE eligible and DEVOTE ineligible populations

|  | No. of patients | No. of events | Person-years | Incidence rate¶<br>(95% CI) | Unadjusted | Adjusted† | Observed Agreement‡ |  |  |
| --- | --- | --- | --- | --- | --- | --- | --- | --- | --- |
|  |  |  |  |  | HR (95% CI) | HR (95% CI§) | SA | EA | SD |
| **CPRD population** |  |  |  |  |  |  |  |  |  |
| Insulin degludec | 812 | 47 | 1,194 | 39.4 (29.6, 52.4) | 0.82 (0.61, 1.10) | 1.36 (0.83, 1.86) | N | N | N |
| Insulin glargine | 9,618 | 910 | 20,001 | 45.5 (42.6, 48.6) | 1.00 (Reference) | 1.00 (Reference) |  |  |  |
| **DEVOTE eligible** |  |  |  |  |  |  |  |  |  |
| Insulin degludec | 376 | 39 | 546 | 71.4 (52.2, 97.8) | 0.86 (0.62, 1.19) | 1.07 (0.63, 1.58) | N | N | Y |
| Insulin glargine | 4,904 | 748 | 9,389 | 79.7 (74.2, 85.6) | 1.00 (Reference) | 1.00 (Reference) |  |  |  |
| **DEVOTE ineligible** |  |  |  |  |  |  |  |  |  |
| Insulin degludec | 436 | 8 | 648 | 12.3 (6.2, 24.7) | 0.77 (0.38, 1.57) | 2.19 (0.30, 3.83) |  |  |  |
| Insulin glargine | 4,714 | 162 | 10,612 | 15.3 (13.1, 17.8) | 1.00 (Reference) | 1.00 (Reference) | N | N | N |
| **DEVOTE Trial** |  |  |  |  |  |  |  |  |  |
| Insulin degludec | 3,818 | 325 | N/A | 42.9 | 0.91 (0.78, 1.06) | N/A |  |  |  |
| Insulin glargine | 3,819 | 356 | N/A | 47.1 | 1.00 (Reference) | N/A |  |  |  |

<sup>¶</sup> Incidence rates are expressed as events per 1,000 person-year. Confidence interval estimated using poisson model.
<sup>†</sup>The following baseline characteristics were included in the propensity score model used for inverse probability of treatment weighting: age, sex, ethnic origin, year of cohort entry, duration of diabetes, BMI, smoking status, A1C level, blood pressure level, eGFR category, comorbidities, antidiabetic drugs, comedications. Multiple imputation was applied for race, Index of Multiple Deprivation decile, smoking status, A1C, eGFR, body mass index, systolic blood pressure, diastolic blood pressure.
<sup>‡</sup>SA: Full statistical significance agreement (Y/N/P) - is said to occur if the RWD and RCT have estimates and CIs on the same side of the null; P: partial significance agreement- met the prespecified noninferiority criteria even though the database study may have indicated superiority, EA: Estimate agreement (Y/N) - is said to occur if the effect estimate of the RWD falls within the 95% confidence interval of the RCT effect estimate. SD: Standardized difference agreement is defined by standardized differences $|Z| < 1.96$ (Y = yes, N = no)
<sup>§</sup>95% CI were bootstrapped using 1000 simulations
S\* suppressed small cells with N < 5
Abbreviations: CI: confidence interval, CPRD: Clinical Practice Research Datalink, HR: hazard ratio, MACE: major adverse cardiovascular event-composite of myocardial infarction, stroke or cardiovascular death,

All the primary analyses failed to reject the null at a p-value of 0.05, which is consistent with the results of the DEVOTE trial. However, due to our wide CIs, we were unable to achieve the FDA non-inferiority upper limit of 1.3 and thus did not achieve statistical agreement with the DEVOTE trial. Estimate agreement was unable to be met as our effect estimates fell outside the CI of the DEVOTE trial. Only the DEVOTE eligible population was able to achieve standardized difference agreement with the trial results.

**Table 4** presents the results of our secondary of the individual components of MACE, HHF, all-cause mortality, and hospitalization due to hypoglycemia. For MI, HR estimates were consistent with findings of the trial across all three observational populations. For stroke, due to low sample size, HR estimates varied greatly. For cardiovascular death, HR estimates were consistent among the RWD populations; however, they were on the opposite side of the null compared to the DEVOTE trial. For HHF, HR estimates were consistent among the three populations. Hospitalization due to hypoglycemia saw differences between the three populations, with the DEVOTE ineligible sub-population effect estimate below the null. For all-cause mortality, the CIs overlapped for the three populations, with all of them achieving standardized difference agreement with the original trial. The overall CPRD population achieved estimate agreement and the DEVOTE eligible achieved full significance agreement with the DEVOTE trial.

**Table 4.**
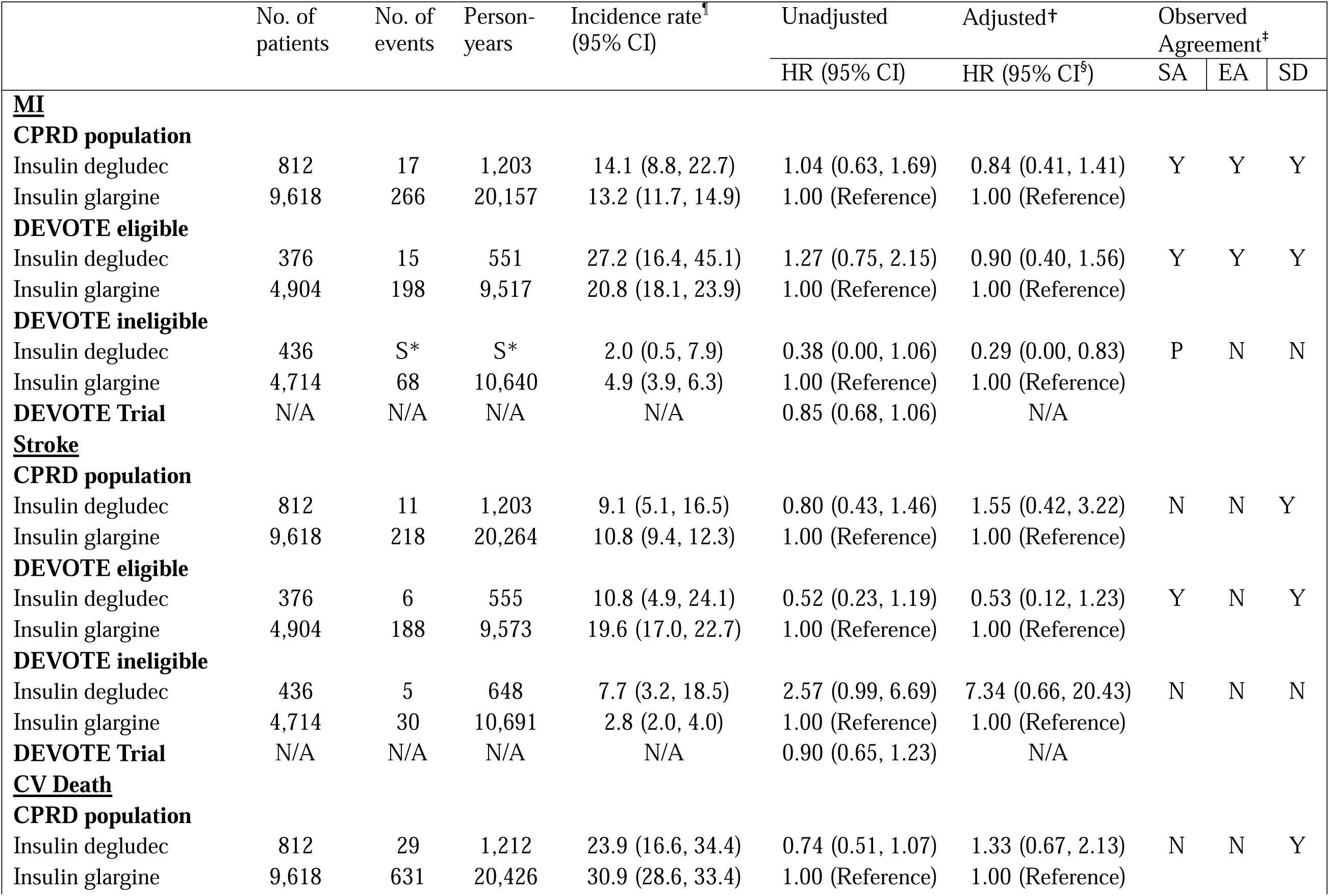
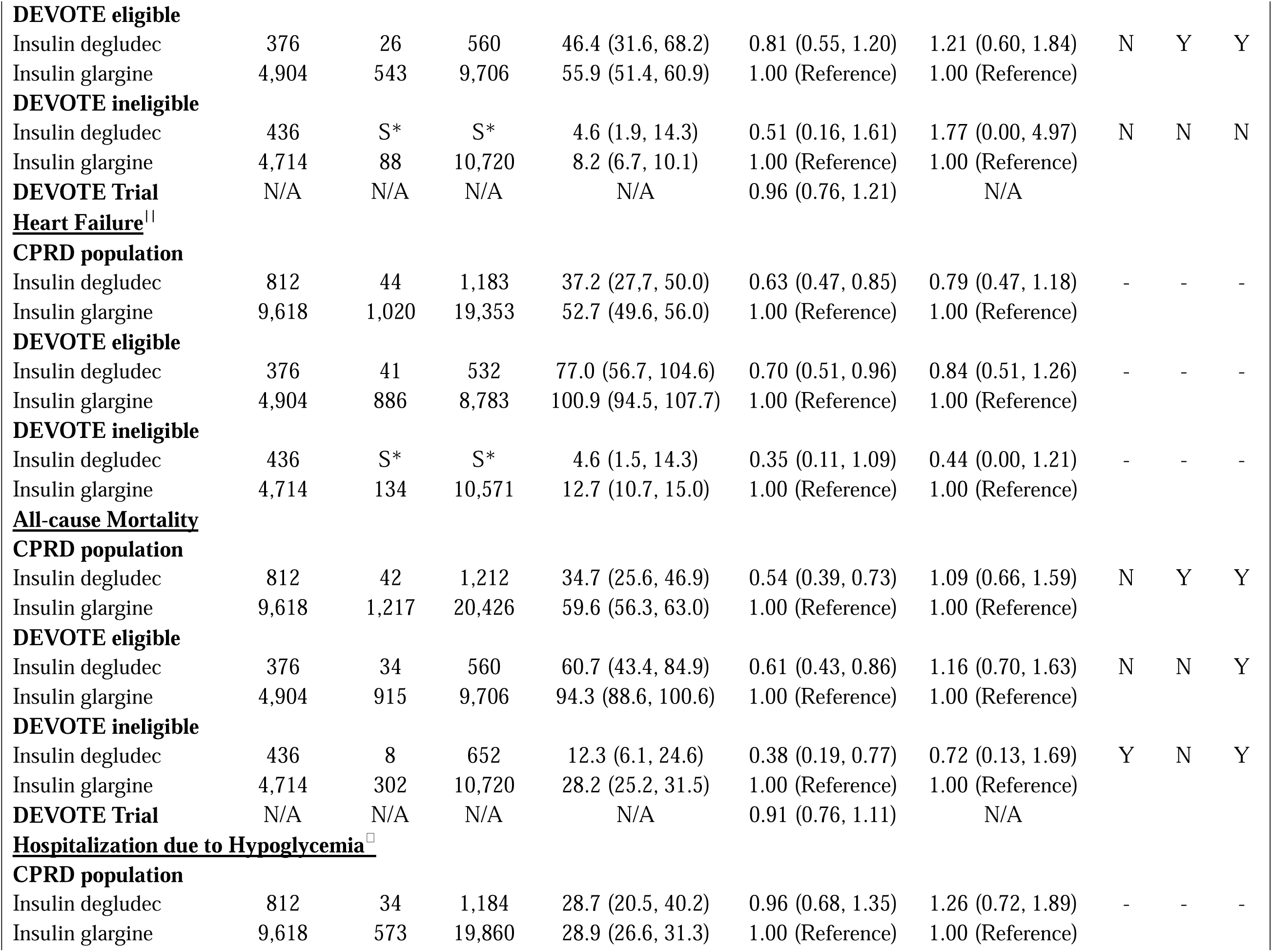
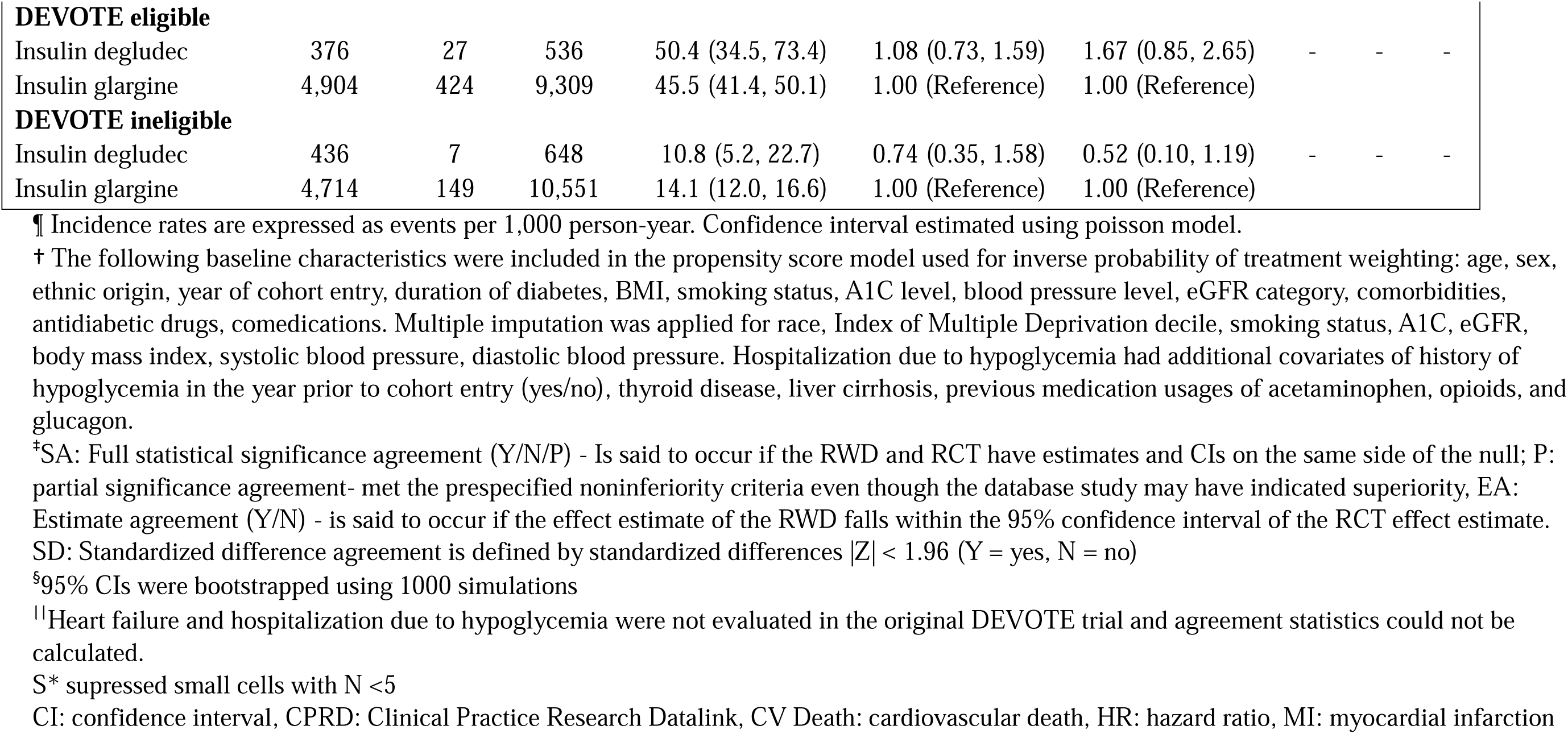
The association between use of insulin degludec compared to insulin glargine for the risk of secondary outcomes including individual components of MACE, heart failure, all-cause mortality and hospitalization due to hypoglycemia, among patients with type 2 diabetes

**Supplemental Table S4** reports the results from our sensitivity analyses including using inverse odds weighting (IOW) to examine transportability. These results from our sensitivity analyses were consistent with those from our primary analysis. **Supplemental Table S5** describe the reasons for censoring among the three populations. Of note, adherence was low in our study, with 80% of our study population discontinuing their treatment before the end of follow-up.

## DISCUSSION

Our study demonstrated that there is a lack of generalizability of the DEVOTE trial to patients with T2DM treated in a real-world setting. Only half of the real-world population would have been eligible for the DEVOTE trial, with notable differences in patient characteristics such as age, use of comedications, and comorbidities between the DEVOTE eligible and ineligible populations. There were also large differences in patient characteristics such as duration of diabetes, comedication use, and comorbidities between the trial and the real-world population in our study. Our findings for the risk of MACE among patients prescribed insulin degludec compared to insulin glargine were consistent across the three populations examined. The DEVOTE eligible subpopulation had an estimate compatible with the original DEVOTE trial, with both suggesting no clinically important difference in risk of MACE between degludec and glargine. In the DEVOTE ineligible population, on the other hand, the low number of events resulted in imprecise estimates. For individual components of MACE, the treatment estimates for MI had the highest degree of agreement with the original DEVOTE trial for all three populations. The other components of MACE, all-cause mortality, HHF, hospitalization due to hypoglycemia had varying levels of consistency among the three populations in the study and, when reported, with the DEVOTE trial. Sensitivity analyses revealed that results were robust to study assumptions.

Trials are known to have limited generalizability to real-world populations, which is evident from our findings. This is, in part, by design. The FDA guidelines for cardiovascular outcome trials target older individuals with T2DM who are at elevated cardiovascular risk which is beneficial in increasing the number of events and reducing the required sample size^20^. Although insulins are exempt from the FDA cardiovascular outcome trial requirement^20^, a similar approach was used in DEVOTE. Due to this trial eligibility criteria, the incidence rate was higher in the DEVOTE eligible population compared to the DEVOTE ineligible or the overall CPRD population in our study. While this selection process resulted in the inclusion of higher-risk patients and a more precise estimate, half of the real-world population was excluded. The use of pragmatic trial designs may increase the generalizability of future trials in this area.

The DEVOTE eligible population often had the highest level of agreement with the DEVOTE trial. This finding provides important reassurance that RWD can be used to complement existing RCT evidence when rigorous methods are used, and attempts are made to emulate RCTs as close as possible. This is especially pertinent to regulators in the US, Canada, and European Union who are incorporating the use of RWD in their decision making^6,7,21^.

We conducted sensitivity analyses to assess transportability such as using IOW to transport our observational study to the trial population based on the distribution of MI in the DEVOTE trial. However, we were limited to aggregate trial-level data. Future research using more granular data will allow for more robust comparisons between patient characteristics of trial and RWD populations using PS and probability distributions^22^. Utilizing patient level trial data for IOW would allow for the inclusion of additional covariates to account for potential effect modifiers and compare effect estimates based on distribution of covariates that more accurately reflect the trial population^23^.

To our knowledge, there have been no previous studies that emulated the DEVOTE trial with RWD. Previous studies have emulated other cardiovascular outcome trials for antidiabetic medications^19,24–26^. For example, the DUPLICATE Team emulated 32 RCTs using non-randomized data from US health care claims data with eight trials evaluating cardiovascular outcomes of antidiabetic medication^19^. All eight emulations achieved either full or partial statistical significance agreement, two did not achieve estimate agreement, and one did not have standardized difference agreement^19^. In a meta-analysis of their 32 trial emulations, they found that the heterogeneity between RCT and RWD effect estimates was mainly attributed to three emulation differences: treatment started in hospital, discontinuation of some baseline treatment at randomization, and delayed onset of drug effects^27^. While our data availability did not include in-hospital drug data and may miss treatments that started in hospital, diabetes treatment often is monitored by the general practitioner in the UK, and we therefore expect subsequent prescriptions to be captured within the CPRD. In terms of discontinuation of some baseline treatment at randomization, the DEVOTE trial allowed for concomitant use of other antidiabetic medications. Our study may be prone to delayed onset of drug effects due to the short duration of medication persistence in clinical practice. Our study adds to this existing body of work as it evaluates an RCT that was not previously studied. It used a primary care database with richer clinical data than administrative claims data.

Our study has many strengths. First, we used a large primary care database that is representative of the real-world population. The CPRD provides clinical laboratory measures such as BMI and A1C to account for clinically relevant covariates. Second, we are the first study to replicate the DEVOTE trial to evaluate the risk of MACE for patients with T2DM prescribed insulin degludec and glargine. Compared to ORIGIN^5^, the other cardiovascular outcome trial in the area, the DEVOTE trial allowed for a stronger emulation. DEVOTE selected for patients who were more likely to receive the treatment in a real-world setting and included an active comparator, reducing time-lag bias and potential confounding by indication. ORIGIN included patients with prediabetes and used standard of care, which is difficult to replicate and may introduce residual confounding. Third, we used IPTW to balance our population and then stabilized and truncated the weights to reduce the influence of extreme weights. Fourth, to our knowledge, we are also the first study emulating cardiovascular outcome trials to compare the patient characteristics and risk of cardiovascular outcomes of the eligible vs non-eligible populations to ascertain differences between those who were and were not eligible to enroll in a trial.

There are several potential limitations to our study. First, due to the small sample size, our study had a low number of events, leading to a lack of precision in our estimates. Second, even after stabilized IPTW weighting, there were some differences in covariate distributions between exposure groups. Residual confounding is thus possible. We did not further adjust for these differences, as we used bootstrapping to adjust our variance, and the current literature recommends against doing so in this setting^28^. Double adjustment only leads to doubly robust estimates if both models are known to be accurate, which we cannot guarantee for our outcome model^28^. Third, given the amount of treatment discontinuation present in our cohort and our use of an ITT approach, exposure misclassification may have biased our results towards the null. Our on-treatment sensitivity analysis saw a shift of our effect estimate towards that reported in the DEVOTE trial.

## Data Availability

All data produced in the present study are available upon reasonable request to the authors

## Funding and Assistance

KBF is supported by a William Dawson Scholar award from McGill University. WW is supported by a McGill Internal Studentship Award and a CANadian Consortium of Clinical Trial TRAINing platform (CANTRAIN) studentship; CANTRAIN is funded by the Canadian Institutes of Health Research

## Conflicts of Interest

None to declare.

## Authors’ contributions

WW led the protocol development and drafted the manuscript. PR conducted the data management and analyses. All authors contributed to protocol development, were involved in data interpretation, critically reviewed the manuscript for important intellectual content, and approved the final manuscript. KBF conceived of the study idea, supervised the study, and is the guarantor.

## Notes

### Competing Interest Statement

The authors have declared no competing interest.

### Author Declarations

The protocol of this study (19_217) was approved by the CPRD's Independent Scientific Advisory Committee (ISAC) and by the CIUSSS West-Central Montreal Research Ethics Board (Montreal, Canada).

