## Supplementary Materials for "Emulating randomized controlled trials of long-acting insulins and cardiovascular events using real-world data for patients with type 2 diabetes"

**Supplemental Methods**

***Definition of agreement statistics:***

*Full statistical significance* *agreement* is said to have occurred if the RWD and RCT have estimates and CIs on the same side of the null. Partial significance agreement is defined as RCT meeting the prespecified noninferiority criteria even though the CPRD study may have indicated superiority^1^. *Estimate agreement* is said to occur if the effect estimate of the RWD falls within the 95% CI of the RCT effect estimate *Standardized difference agreement* is defined by standardized differences |Z| < 1.96.

Z =$\frac{\hat{\theta}_{\mathrm{RWE}}-\hat{\theta}_{\mathrm{RCT}}}{\sqrt{{\hat{\sigma}^{2}}_{\mathrm{RWE}}+{\hat{\sigma}^{2}}_{\mathrm{RCT}}}}$ $\hat{\theta}$ are effect estimates and $\hat{\sigma}^{2}$ are variances.

As per the FDA, all the major CVOTs were designed for non-inferiority with an upper CI limit of 1.3^2^. For our statistical significance agreement statistic, we first assessed whether the trial was able to demonstrate non-inferiority and then evaluated whether the RWD study was able to replicate the non-inferiority finding within the same margin. If the trial achieved non-inferiority but the RWD study achieved superiority, partial statistical significance agreement was established. In addition to non-inferiority, if superiority was established in the trial, we assessed if the RWD study also found superiority to achieve full statistical significance agreement.

***Sensitivity analyses:***

We conducted 6 different sensitivity analyses. First, to compare our subpopulations to the trial ones, we combined inverse odds weights (IOW) targeting the distribution of prior MI in DEVOTE trial with our stabilized IPTW^3^. In this analysis, we reweighed our three study populations to mirror the distribution of MI in the DEVOTE trial (34%)^4^. Second, we explored the use of applying IOW to the DEVOTE eligible population to target the DEVOTE ineligible population rather than the DEVOTE trial. These IOW included all the covariates initially used in our IPTW model rather than only MI. Third, as trials usually are shorter in duration compared to observational studies, we repeated our primary analysis with follow-up time restricted to a maximum of 2 years to replicate the follow-up duration of the DEVOTE trial. Fourth, to examine the impact of exposure discontinuation and switching, we described the rate and timing of exposure discontinuation and switching. Fifth, we repeated our primary analysis using an ‘on treatment’ exposure definition in which we censored patients who discontinued or switched treatments. Sixth, we matched our exposure groups based on age, year of cohort entry, and the logit of the PS with a caliper at 0.05 and re-ran our primary analysis.

**Supplemental Table S1** Operationalization of DEVOTE trial inclusion criteria for observational study and number of patients for each corresponding criteria

| **RCT inclusion Criteria** | **Observational Study Operationalization of Inclusion Criteria** | **Number of patients** |
| --- | --- | --- |
| Criteria 1: Age ≥ 50 years old at cohort entry and at least 1 of the following conditions: |  | 4,826 |
| 1. Prior myocardial infarction | 1. Code for MI in CPRD or HES before cohort entry | 2,280 |
| 2. Prior stroke or prior Transient Ischemic Attack (TIA) | 2. Codes for stroke or TIA in CPRD or HES before cohort entry | 1,188 |
| 3. Prior coronary, carotid, or peripheral arterial revascularization | 3. Coronary, carotid or peripheral arterial revascularization code in CPRD or HES before cohort entry | 938 |
| 4. >50% stenosis on angiography or other imaging of coronary, carotid or lower-extremity artery | 4. Stenosis of coronary, carotid or lower-extremity artery codes in CPRD or HES before cohort entry | 118 |
| 5. History of symptomatic coronary heart disease documented by positive exercise stress test of any cardiac imaging, or unstable angina pectoris with ECG changes | 5. Coronary artery disease codes in CPRD or HES before cohort entry | 2,917 |
| 6. Chronic heart failure NYHA class II-III | 6. Code for heart failure in CPRD or HES before cohort entry | 1,523 |
| 7. Chronic kidney disease corresponding to estimated glomerular filtration rate (eGFR) of 30-59 mL/min per 1.73m^2 per CKD-EPI | 7. Codes for eGFR 30-59 mL/min per 1.73m^2 or codes for stage 3 CKD in CPRD or HES before cohort entry | 2,441 |
| Criteria 2: Age ≥ 60 years old at cohort entry and at least 1 of the following risk factors: |  | 2,922 |
| 1. Microalbuminuria or proteinuria | 1. Codes for microalbuminuria or proteinuria in CPRD or HES before cohort entry | 2,102 |
| 2. Hypertension and left ventricular hypertrophy by ECG or imaging | 2. Codes for hypertension or left ventricular hypertrophy in CPRD or HES before cohort entry | 489 |
| 3. Left ventricular systolic and diastolic dysfunction by imaging | 3. Codes for left ventricular systolic and diastolic dysfunction in CPRD or HES before cohort entry | 19 |
| 4. Ankle/brachial index <0.9 | 4. Codes for peripheral vascular disease in CPRD or HES before cohort entry | 1,023 |
| Total participants that match Criteria 1 or 2 | | **5,470** |
| Include patients with A1C ≥ 7.0% or current insulin treatment corresponding to ≥ 20 U/d of basal insulin | A1C ≥ 7.0% or prescription of basal insulin (NPH insulin or insulin detemir) before cohort entry | 5,338 (132 excluded from above) |
| One or more oral or injectable antidiabetic agent(s) | Prescription of metformin, sulfonylureas, thiazolidinediones, DPP-4 inhibitor, GLP-1 RA, alpha-glucosidase inhibitors, meglitinides, SGLT-2 inhibitors, insulin (except insulin glargine or insulin degludec) before cohort entry | 5,280 (58 excluded from above) |
| Total DEVOTE eligible population |  | **5,280** |

Abbreviations: A1C: glycated hemoglobin, CKD-EPI: Chronic Kidney Disease Epidemiology Collaboration, CPRD: Clinical Practice Research Datalink, DPP-4 inhibitors: dipeptidyl peptidase-4 inhibitors, ECG: electrocardiogram, eGFR: estimated glomerular filtration rate, GLP-1 RA: Glucagon-like peptide-1 receptor agonists, HES: Hospital Episode Statistics, U/d: units per day, MI: myocardial infarction, NYHA: New York Heart Association, SGLT-2 inhibitors: sodium-glucose cotransporter-2 inhibitors, TIA: transient ischemic stroke

^¶^ ICD-10 codes for HES

**Supplemental Table S2** Baseline characteristics of CPRD, DEVOTE eligible and DEVOTE ineligible populations of patients with type 2 diabetes who initiated insulin degludec or insulin glargine before inverse probability of treatment weighting and multiple imputation.

|  | CPRD | | | DEVOTE eligible | | | DEVOTE ineligible | | |
| --- | --- | --- | --- | --- | --- | --- | --- | --- | --- |
|  | Insulin glargine users | Insulin degludec users | SMD | Insulin glargine users | Insulin degludec users | SMD | Insulin glargine users | Insulin degludec users | SMD |
|  | N/mean  (%/SD) | N/mean (%/SD) |  | N/mean  (%/SD) | N/mean (%/SD) |  | N/mean  (%/SD) | N/mean (%/SD) |  |
| Number of patients | 9,618 | 812 |  | 4,904 | 376 |  | 4,714 | 436 |  |
| *Ethnicity- Missing* | *775 (8.1)* | *78 (9.6)* | *0.05* | *173 (3.5)* | *16 (4.3)* | *0.04* | *602 (12.8)* | *62 (14.2)* | *0.04* |
| Age group, n (%) |  |  |  |  |  |  |  |  |  |
| <49.9 | 1,562 (16.2) | 145 (17.9) | 0.04 | 0 (0.0) | 0 (0.0) | **---** | 1,562 (33.1) | 145 (33.3) | 0.00 |
| 50-59.9 | 2,166 (22.5) | 247 (30.4) | 0.18 | 632 (12.9) | 78 (20.7) | 0.21 | 1,534 (32.5) | 169 (38.8) | 0.13 |
| 60-69.9 | 2,299 (23.9) | 222 (27.3) | 0.08 | 1,414 (28.8) | 144 (38.3) | 0.20 | 885 (18.8) | 78 (17.9) | 0.02 |
| 70-79.9 | 2,126 (22.1) | 135 (16.6) | 0.14 | 1,593 (32.5) | 99 (26.3) | 0.14 | 533 (11.3) | 36 (8.3) | 0.10 |
| 80+ | 1,465 (15.2) | 63 (7.8) | 0.24 | 1,265 (25.8) | 55 (14.6) | 0.28 | 200 (4.2) | 8 (1.8) | 0.14 |
| Index of Multiple Deprivation, n(%) |  |  |  |  |  |  |  |  |  |
| 1 | 1,612 (16.8) | 178 (21.9) | 0.13 | 869 (17.7) | 90 (23.9) | 0.15 | 743 (15.8) | 88 (20.2) | 0.12 |
| 2 | 1,797 (18.7) | 159 (19.6) | 0.02 | 947 (19.3) | 68 (18.1) | 0.03 | 850 (18.0) | 91 (20.9) | 0.07 |
| 3 | 1,867 (19.4) | 149 (18.3) | 0.03 | 958 (19.5) | 75 (19.9) | 0.01 | 909 (19.3) | 74 (17.0) | 0.06 |
| 4 | 2,065 (21.5) | 163 (20.1) | 0.03 | 1,014 (20.7) | 72 (19.1) | 0.04 | 1,051 (22.3) | 91 (20.9) | 0.03 |
| 5 | 2,271 (23.6) | 163 (20.1) | 0.09 | 1,112 (22.7) | 71 (18.9) | 0.09 | 1,159 (24.6) | 92 (21.1) | 0.08 |
| *Missing* | *6 (0.1)* | *S** | *-* | *4 (0.1)* | *S** | *-* | *S** | *S** | *-* |
| Lifestyle and other covariates |  |  |  |  |  |  |  |  |  |
| BMI |  |  |  |  |  |  |  |  |  |
| < 25 | 1,756 (18.3) | 92 (11.3) | 0.20 | 890 (18.1) | 48 (12.8) | 0.15 | 866 (18.4) | 44 (10.1) | 0.25 |
| 25-29.9 | 2,843 (29.6) | 183 (22.5) | 0.17 | 1,526 (31.1) | 76 (20.2) | 0.26 | 1,317 (27.9) | 107 (24.5) | 0.09 |
| 30.0-34.9 | 2,500 (26.0) | 217 (26.7) | 0.01 | 1,286 (26.2) | 97 (25.8) | 0.01 | 1,214 (25.8) | 120 (27.5) | 0.03 |
| 35-39.9 | 1,338 (13.9) | 169 (20.8) | 0.18 | 689 (14.0) | 86 (22.9) | 0.23 | 649 (13.8) | 83 (19.0) | 0.13 |
| 40+ | 935 (9.7) | 143 (17.6) | 0.23 | 427 (8.7) | 65 (17.3) | 0.26 | 508 (10.8) | 78 (17.9) | 0.20 |
| *Missing* | *246 (2.6)* | *8 (1.0)* | *0.12* | *86 (1.8)* | *S** | *-* | *160 (3.4)* | *S** | *-* |
| *Smoking- Missing* | *157 (1.6)* | 7 (0.9) | *0.07* | *57 (1.2)* | *3 (0.8)* | *0.04* | *100 (2.1)* | *S** | *-* |
| *SBP- Missing* | *43 (0.4)* | *S** | *-* | *S** | *S** | *-* | *42 (0.9)* | *S** | *-* |
| *DBP- Missing* | *43 (0.4)* | *S** | *-* | *S** | *S** | *-* | *42 (0.9)* | *S** | *-* |
| eGFR, ml/min/1.73m2 |  |  |  |  |  |  |  |  |  |
| <60 | 2,897 (30.1) | 166 (20.4) | 0.24 | 2,510 (51.2) | 140 (37.2) | 0.28 | 387 (8.2) | 26 (6.0) | 0.10 |
| 60+ | 6,491 (67.5) | 638 (78.6) |  | 2,366 (48.2) | 233 (62.0) |  | 4,125 (87.5) | 405 (92.9) |  |
| *Missing* | *230 (2.4)* | *8 (1.0)* | *0.11* | *28 (0.6)* | *S** | *-* | *202 (4.3)* | *5 (1.1)* | *0.19* |
| A1C |  |  |  |  |  |  |  |  |  |
| ≤ 7 % | 765 (8.0) | 43 (5.3) | 0.11 | 421 (8.6) | 15 (4.0) | 0.19 | 344 (7.3) | 28 (6.4) | 0.04 |
| 7.1-8.0% | 1,186 (12.3) | 96 (11.8) | 0.02 | 706 (14.4) | 46 (12.2) | 0.06 | 480 (10.2) | 50 (11.5) | 0.03 |
| > 8.0% | 7,477 (77.7) | 670 (82.5) | 0.09 | 3,774 (77.0) | 315 (83.8) | 0.17 | 3,703 (78.6) | 355 (81.4) | 0.00 |
| *Missing* | *190 (2.0)* | *S** | *-* | *S** | *S** | *-* | *187 (4.0)* | *S** | *-* |
| Comedications, n (%) |  |  |  |  |  |  |  |  |  |
| ACE inhibitors | 4,375 (45.5) | 400 (49.3) | 0.08 | 2,594 (52.9) | 205 (54.5) | 0.03 | 1,781 (37.8) | 195 (44.7) | 0.14 |
| Acetylsalicylic acid | 2,982 (31.0) | 225 (27.7) | 0.07 | 2,196 (44.8) | 154 (41.0) | 0.08 | 786 (16.7) | 71 (16.3) | 0.01 |
| Antiplatelets | 3,522 (36.6) | 269 (33.1) | 0.07 | 2,666 (54.4) | 194 (51.6) | 0.06 | 856 (18.2) | 75 (17.2) | 0.03 |
| Angiotensin II receptor blockers | 1,773 (18.4) | 159 (19.6) | 0.03 | 1,190 (24.3) | 98 (26.1) | 0.04 | 583 (12.4) | 61 (14.0) | 0.05 |
| Beta-blockers | 3,029 (31.5) | 227 (28.0) | 0.08 | 2,348 (47.9) | 168 (44.7) | 0.06 | 681 (14.4) | 59 (13.5) | 0.03 |
| Calcium-channel blockers | 3,005 (31.2) | 245 (30.2) | 0.02 | 1,953 (39.8) | 147 (39.1) | 0.01 | 1,052 (22.3) | 98 (22.5) | 0.00 |
| Diuretics | 3,260 (33.9) | 247 (30.4) | 0.07 | 2,389 (48.7) | 178 (47.3) | 0.03 | 871 (18.5) | 69 (15.8) | 0.07 |
| Fibrates | 286 (3.0) | 23 (2.8) | 0.01 | 173 (3.5) | 7 (1.9) | 0.10 | 113 (2.4) | 16 (3.7) | 0.07 |
| Oral anticoagulants | 1,029 (10.7) | 72 (8.9) | 0.06 | 885 (18.0) | 60 (16.0) | 0.06 | 144 (3.1) | 12 (2.8) | 0.02 |
| Nonsteroidal anti-inflammatory drugs | 2,421 (25.2) | 222 (27.3) | 0.05 | 1,238 (25.2) | 111 (29.5) | 0.10 | 1,183 (25.1) | 111 (25.5) | 0.01 |
| Statins | 7,309 (76.0) | 642 (79.1) | 0.07 | 4,109 (83.8) | 326 (86.7) | 0.08 | 3,200 (67.9) | 316 (72.5) | 0.10 |
| Hypoglycemia specific covariates |  |  |  |  |  |  |  |  |  |
| Thyroid disease | 1,357 (14.1) | 115 (14.2) | 0.00 | 836 (17.0) | 66 (17.6) | 0.01 | 521 (11.1) | 49 (11.2) | 0.01 |
| Acetaminophen | 4,177 (43.4) | 328 (40.4) | 0.06 | 2,536 (51.7) | 189 (50.3) | 0.03 | 1,641 (34.8) | 139 (31.9) | 0.06 |
| Opioids | 3,962 (41.2) | 344 (42.4) | 0.02 | 2,228 (45.4) | 181 (48.1) | 0.05 | 1,734 (36.8) | 163 (37.4) | 0.01 |
| Glucagon | 78 (0.8) | 7 (0.9) | 0.01 | 32 (0.7) | 5 (1.3) | 0.07 | 46 (1.0) | S* | - |

S* supressed small cells with N <5 Abbreviations: A1C: glycated hemoglobin, ACE inhibitors: angiotensin-converting enzyme inhibitors, BMI: body mass index, DBP: diastolic blood pressure, CPRD: Clinical Practice Research Datalink, eGFR: estimated glomerular filtration rate, SBP: systolic blood pressure, SD: standard deviation, SMD: standardized mean difference

**Supplemental Table S3** Baseline characteristics of CPRD, DEVOTE eligible and DEVOTE ineligible populations of patients with type 2 diabetes who initiated insulin degludec or insulin glargine after inverse probability of treatment weighting and multiple imputation.

|  | CPRD | | | DEVOTE eligible | | | DEVOTE ineligible | | |
| --- | --- | --- | --- | --- | --- | --- | --- | --- | --- |
|  | Insulin glargine users | Insulin degludec users | SMD | Insulin glargine users | Insulin degludec users | SMD | Insulin glargine users | Insulin degludec users | SMD |
|  | N/mean  (%/SD) | N/mean (%/SD) |  | N/mean  (%/SD) | N/mean (%/SD) |  | N/mean  (%/SD) | N/mean (%/SD) |  |
| Number of patients | 9,637 | 769 |  | 4,916 | 320 |  | 4,722 | 414 |  |
| *Ethnicity- Missing* | *0 (0.0)* | *0 (0.0)* | *---* | *0 (0.0)* | *0 (0.0)* | *---* | *0 (0.0)* | *0 (0.0)* | *---* |
| Age group, n (%) |  |  |  |  |  |  |  |  |  |
| <49.9 | 1,576 (16.4) | 110 (14.3) | 0.06 | 0 (0.0) | 0 (0.0) | **---** | 1,565 (33.1) | 118 (28.5) | 0.10 |
| 50-59.9 | 2,234 (23.2) | 216 (28.1) | 0.11 | 662 (13.5) | 48 (14.9) | 0.04 | 1,562 (33.1) | 160 (38.6) | 0.12 |
| 60-69.9 | 2,333 (24.2) | 173 (22.5) | 0.04 | 1,458 (29.7) | 95 (29.5) | 0.00 | 883 (18.7) | 69 (16.7) | 0.05 |
| 70-79.9 | 2,085 (21.6) | 188 (24.4) | 0.07 | 1,571 (31.9) | 120 (37.5) | 0.12 | 521 (11.0) | 46 (11.1) | 0.00 |
| 80+ | 1,408 (14.6) | 82 (10.6) | 0.12 | 1,225 (24.9) | 58 (18.1) | 0.17 | 191 (4.0) | 21 (5.1) | 0.05 |
| Index of Multiple Deprivation, n(%) |  |  |  |  |  |  |  |  |  |
| 1 | 1,661 (17.2) | 155 (20.2) | 0.08 | 898 (18.3) | 72 (22.6) | 0.11 | 764 (16.2) | 81 (19.5) | 0.09 |
| 2 | 1,808 (18.8) | 143 (18.6) | 0.00 | 945 (19.2) | 55 (17.1) | 0.06 | 865 (18.3) | 87 (20.9) | 0.07 |
| 3 | 1,863 (19.3) | 147 (19.1) | 0.01 | 960 (19.5) | 69 (21.6) | 0.05 | 901 (19.1) | 65 (15.7) | 0.09 |
| 4 | 2,060 (21.4) | 176 (22.9) | 0.04 | 1,013 (20.6) | 62 (19.5) | 0.03 | 1,047 (22.2) | 100 (24.1) | 0.04 |
| 5 | 2,244 (23.3) | 147 (19.1) | 0.10 | 1,099 (22.4) | 62 (19.3) | 0.08 | 1,146 (24.3) | 82 (19.8) | 0.11 |
| *Missing* | *0 (0.0)* | *0 (0.0)* | **---** | *0 (0.0)* | *0 (0.0)* | --- | *0 (0.0)* | *0 (0.0)* | --- |
| Lifestyle and other covariates |  |  |  |  |  |  |  |  |  |
| BMI |  |  |  |  |  |  |  |  |  |
| < 25 | 1,753 (18.2) | 113 (14.8) | 0.09 | 887 (18.0) | 45 (14.0) | 0.11 | 858 (18.2) | 63 (15.3) | 0.08 |
| 25-29.9 | 2,855 (29.6) | 235 (30.6) | 0.02 | 1,515 (30.8) | 88 (27.4) | 0.08 | 1,366 (28.9) | 138 (33.2) | 0.09 |
| 30.0-34.9 | 2,580 (26.8) | 222 (28.8) | 0.05 | 1,314 (26.7) | 102 (32.0) | 0.12 | 1,260 (26.7) | 111 (26.8) | 0.00 |
| 35-39.9 | 1,431 (14.8) | 113 (14.6) | 0.01 | 740 (15.1) | 52 (16.2) | 0.03 | 692 (14.7) | 52 (12.5) | 0.06 |
| 40+ | 1,018 (10.6) | 86 (11.2) | 0.02 | 460 (9.4) | 34 (10.5) | 0.04 | 547 (11.6) | 51 (12.2) | 0.02 |
| *Missing* | *0 (0.0)* | *0 (0.0)* | *---* | *0 (0.0)* | *0 (0.0)* | *---* | *0 (0.0)* | *0 (0.0)* | *---* |
| *Smoking- Missing* | *0 (0.0)* | *0 (0.0)* | *---* | *0 (0.0)* | *0 (0.0)* | *---* | *0 (0.0)* | *0 (0.0)* | *---* |
| *SBP- Missing* | *0 (0.0)* | *0 (0.0)* | *---* | *0 (0.0)* | *0 (0.0)* | *---* | *0 (0.0)* | *0 (0.0)* | *---* |
| *DBP- Missing* | *0 (0.0)* | *0 (0.0)* | *---* | *0 (0.0)* | *0 (0.0)* | *---* | *0 (0.0)* | *0 (0.0)* | *---* |
| eGFR, ml/min/1.73m2 |  |  |  |  |  |  |  |  |  |
| <60 | 2,859 (29.7) | 217 (28.2) | 0.03 | 2,468 (50.2) | 158 (49.3) | 0.02 | 395 (8.4) | 29 (7.0) | 0.05 |
| 60+ | 6,778 (70.3) | 552 (71.8) |  | 2,448 (49.8) | 163 (50.7) |  | 4,327 (91.6) | 385 (93.0) | 0.00 |
| *Missing* | *0 (0.0)* | *0 (0.0)* | *---* | *0 (0.0)* | *0 (0.0)* | *---* | *0 (0.0)* | *0 (0.0)* | *---* |
| HbA1c |  |  |  |  |  |  |  |  |  |
| ≤ 7 % | 758 (7.9) | 40 (5.2) | 0.11 | 404 (8.2) | 17 (5.4) | 0.11 | 361 (7.6) | 22 (5.3) | 0.10 |
| 7.1-8.0% | 1,197 (12.4) | 82 (10.7) | 0.05 | 700 (14.2) | 52 (16.3) | 0.06 | 500 (10.6) | 29 (7.1) | 0.12 |
| > 8.0% | 7,681 (79.7) | 647 (84.1) | 0.12 | 3,811 (77.5) | 251 (78.2) | 0.02 | 3,861 (81.8) | 363 (87.7) | 0.16 |
| *Missing* | *0 (0.0)* | *0 (0.0)* | *---* | *0 (0.0)* | *0 (0.0)* | *---* | *0 (0.0)* | *0 (0.0)* | *---* |
| Comedications, n (%) |  |  |  |  |  |  |  |  |  |
| ACE inhibitors | 4,411 (45.8) | 364 (47.4) | 0.03 | 2,608 (53.1) | 172 (53.6) | 0.01 | 1,809 (38.3) | 168 (40.4) | 0.04 |
| Acetylsalicylic acid | 2,961 (30.7) | 257 (33.3) | 0.06 | 2,186 (44.5) | 149 (46.3) | 0.04 | 785 (16.6) | 80 (19.3) | 0.07 |
| Antiplatelets | 3,500 (36.3) | 305 (39.7) | 0.07 | 2,661 (54.1) | 183 (57.2) | 0.06 | 854 (18.1) | 95 (22.8) | 0.12 |
| Angiotensin II receptor blockers | 1,788 (18.6) | 133 (17.3) | 0.03 | 1,197 (24.4) | 69 (21.6) | 0.07 | 595 (12.6) | 53 (12.8) | 0.01 |
| Beta-blockers | 3,007 (31.2) | 227 (29.5) | 0.04 | 2,342 (47.6) | 147 (46.0) | 0.03 | 677 (14.3) | 47 (11.4) | 0.09 |
| Calcium-channel blockers | 2,999 (31.1) | 216 (28.0) | 0.07 | 1,953 (39.7) | 123 (38.4) | 0.03 | 1,054 (22.3) | 76 (18.5) | 0.10 |
| Diuretics | 3,239 (33.6) | 254 (33.0) | 0.01 | 2,388 (48.6) | 161 (50.2) | 0.03 | 864 (18.3) | 69 (16.8) | 0.04 |
| Fibrates | 285 (3.0) | 26 (3.3) | 0.02 | 168 (3.4) | 17 (5.4) | 0.10 | 117 (2.5) | 11 (2.6) | 0.01 |
| Oral anticoagulants | 1,016 (10.5) | 84 (10.9) | 0.01 | 878 (17.9) | 57 (17.8) | 0.00 | 143 (3.0) | 16 (4.0) | 0.05 |
| Nonsteroidal anti-inflammatory drugs | 2,442 (25.3) | 186 (24.2) | 0.03 | 1,254 (25.5) | 78 (24.4) | 0.03 | 1,189 (25.2) | 100 (24.2) | 0.02 |
| Statins | 7,348 (76.2) | 583 (75.8) | 0.01 | 4,129 (84.0) | 267 (83.3) | 0.02 | 3,226 (68.3) | 279 (67.3) | 0.02 |
| Hypoglycemia specific covariates |  |  |  |  |  |  |  |  |  |
| Thyroid disease | 1,359 (14.1) | 123 (16.0) | 0.05 | 839 (17.1) | 70 (22.0) | 0.12 | 524 (11.1) | 49 (11.9) | 0.03 |
| Acetaminophen | 4,167 (43.2) | 315 (41.0) | 0.05 | 2,536 (51.6) | 172 (53.6) | 0.04 | 1,638 (34.7) | 121 (29.1) | 0.12 |
| Opioids | 3,977 (41.3) | 336 (43.7) | 0.05 | 2,242 (45.6) | 166 (51.7) | 0.12 | 1,739 (36.8) | 152 (36.8) | 0.00 |
| Glucagon | 76 (0.8) | 15 (2.0) | 0.10 | 32 (0.6) | 14 (4.2) | 0.23 | 45 (0.9) | *S** | *-* |

Abbreviations: A1C: glycated hemoglobin, ACE inhibitors: angiotensin-converting enzyme inhibitors, BMI: body mass index, DBP: diastolic blood pressure, CPRD: Clinical Practice Research Datalink, eGFR: estimated glomerular filtration rate, SBP: systolic blood pressure, SD: standard deviation, SMD: standardized mean difference

**Supplemental Table S4** Sensitivity analyses for the association of insulin degludec compared to insulin glargine and risk of MACE among patients with type 2 diabetes

|  | | No. of patients | No. of events | Person-years | Incidence rate* (95% CI) | Unadjusted | Adjusted† |
| --- | --- | --- | --- | --- | --- | --- | --- |
|  |  |  |  |  |  | HR (95% CI) | HR (95% CI^‡^) |
| **2 Year Follow-up**  **CPRD population** | | | |  |  |  |  |
| Insulin degludec | 812 | | 38 | 965 | 39.4 (28.6, 54.1) | 0.77 (0.56, 1.07) | 1.36 (0.82, 2.27) |
| Insulin glargine | 9,618 | | 672 | 13,336 | 50.4 (46.7, 54.3) | 1.00 (Reference) | 1.00 (Reference) |
| **DEVOTE eligible** |  | |  |  |  |  |  |
| Insulin degludec | 376 | | 32 | 446 | 71.7 (50.7, 101.4) | 0.83 (0.58, 1.18) | 0.91 (0.57, 1.47) |
| Insulin glargine | 4,904 | | 557 | 6,468 | 86.1 (79.3, 93.6) | 1.00 (Reference) | 1.00 (Reference) |
| **DEVOTE ineligible** |  | |  |  |  |  |  |
| Insulin degludec | 436 | | 6 | 519 | 11.6 (5.2, 25.7) | 0.68 (0.30, 1.54) | 2.67 (0.90, 7.91) |
| Insulin glargine | 4,714 | | 115 | 6,868 | 16.7 (13.9, 20.1) | 1.00 (Reference) | 1.00 (Reference) |
| **‘On-treatment’**^§^  **CPRD population** | | |  |  |  |  |  |
| Insulin degludec | 812 | | 14 | 462 | 30.3 (18.0, 51.2) | 0.62 (0.36, 1.07) | 1.41 (0.60, 3.29) |
| Insulin glargine | 9,618 | | 229 | 4,490 | 51.0 (44.8, 58.1) | 1.00 (Reference) | 1.00 (Reference) |
| **DEVOTE eligible** |  | |  |  |  |  |  |
| Insulin degludec | 376 | | 12 | 227 | 52.9 (30.0, 93.2) | 0.68 (0.38, 1.22) | 0.87 (0.43, 1.74) |
| Insulin glargine | 4,904 | | 190 | 2,237 | 84.9 (73.7, 97.9) | 1.00 (Reference) | 1.00 (Reference) |
| **DEVOTE ineligible** |  | |  |  |  |  |  |
| Insulin degludec | 436 | | *S** | *S** | 8.5 (2.1, 34.1) | 0.48 (0.12, 1.99) | 3.23 (1.65, 6.31) |
| Insulin glargine | 4,714 | | 39 | 2,253 | 17.3 (12.6, 23.7) | 1.00 (Reference) | 1.00 (Reference) |
| **Propensity Score Matching**^\|\|^  **CPRD population** | | |  |  |  |  |  |
| Insulin degludec | 769 | | 35 | 1,102 | 31.8 (22.8, 44.2) | 1.46 (0.90, 2.38) | 1.18 (0.63, 2.20) |
| Insulin glargine | 769 | | 47 | 1,110 | 42.3 (31.8, 56.3) | 1.00 (Reference) | 1.00 (Reference) |
| **DEVOTE eligible** |  | |  |  |  |  |  |
| Insulin degludec | 332 | | 36 | 454 | 79.3 (57.2, 109.9) | 1.14 (0.63, 2.05) | 1.10 (0.50, 2.38) |
| Insulin glargine | 332 | | 37 | 456 | 81.1 (58.8, 112.0) | 1.00 (Reference) | 1.00 (Reference) |
| **DEVOTE ineligible** |  | |  |  |  |  |  |
| Insulin degludec | 402 | | 6 | 589 | 10.2 (4.6, 22.7) | 1.05 (0.30, 3.61) | ---^¶^ |
| Insulin glargine | 402 | | 6 | 612 | 9.8 (4.4, 22.7) | 1.00 (Reference) | 1.00 (Reference) |
| **IOW with MI**^#^  **Overall CPRD population** | | |  |  |  |  |  |
| Insulin degludec | 812 | | 47 | 1,194 | 39.4 (29.6, 52.4) | 0.82 (0.61, 1.10) | 1.35 (0.87, 2.08) |
| Insulin glargine | 9,618 | | 910 | 20,001 | 45.5 (42.6, 48.6) | 1.00 (Reference) | 1.00 (Reference) |
| **DEVOTE eligible** |  | |  |  |  |  |  |
| Insulin degludec | 376 | | 39 | 546 | 71.4 (52.2, 97.8) | 0.86 (0.62, 1.19) | 1.07 (0.69, 1.67) |
| Insulin glargine | 4,904 | | 748 | 9,389 | 79.7 (74.2, 85.6) | 1.00 (Reference) | 1.00 (Reference) |
| **DEVOTE ineligible** |  | |  |  |  |  |  |
| Insulin degludec | 436 | | 8 | 648 | 12.3 (6.2, 24.7) | 0.77 (0.38, 1.57) | 0.78 (0.28, 2.20) |
| Insulin glargine | 4,714 | | 162 | 10,612 | 15.3 (13.1, 17.8) | 1.00 (Reference) | 1.00 (Reference) |
| **IOW with DEVOTE ineligible**** | | |  |  |  |  |  |
| **DEVOTE eligible** |  | |  |  |  |  |  |
| Insulin degludec | 376 | | 39 | 546 | 71.4 (52.2, 97.8) | 0.86 (0.62, 1.19) | 1.07 (0.57, 2.01) |
| Insulin glargine | 4,904 | | 748 | 9,389 | 79.7 (74.2, 85.6) | 1.00 (Reference) | 1.00 (Reference) |

*Incidence rates are expressed as events per 1,000 person-year. Confidence interval estimated using Poisson model.

†The following baseline characteristics were included in the propensity score model used for inverse probability of treatment weighting: age, sex, ethnic origin, year of cohort entry, duration of diabetes, BMI, smoking status, A1C level, blood pressure level, eGFR category, comorbidities, antidiabetic drugs, comedications. Multiple imputation was applied for race, Index of Multiple Deprivation decile, smoking status, A1C, eGFR, body mass index, systolic blood pressure, diastolic blood pressure.

^‡^95% CIs were estimated using robust sandwich variance estimator.

^¶^ Estimate was unable to converge

S* supressed small cells with N <5

^§^On-treatment analysis where patients were censored for treatment discontinuation or switching

^||^Matching analysis where we matched our exposure groups based on age, year of cohort entry and the logit of the PS with a caliper at 0.05

^#^Inverse odds weighting analysis where we inverse odds weights targeting the distribution of prior MI in DEVOTE trial participants (34%) with our stabilized and truncated IPTW

**Inverse odds weighting to the DEVOTE eligible population to target the DEVOTE ineligible. These IOW included al the covariates initially used in our IPTW model

Abbreviations: CI: confidence interval, CPRD: Clinical Practice Research Datalink, IOW: inverse odds weights, IPTW: inverse probability of treatment weighting, MI: myocardial infarction

**Supplemental Figure S2** Plot of absolute standardized mean differences for patient characteristics in total DEVOTE eligible and DEVOTE ineligible populations prior to IPTW weighting and imputation

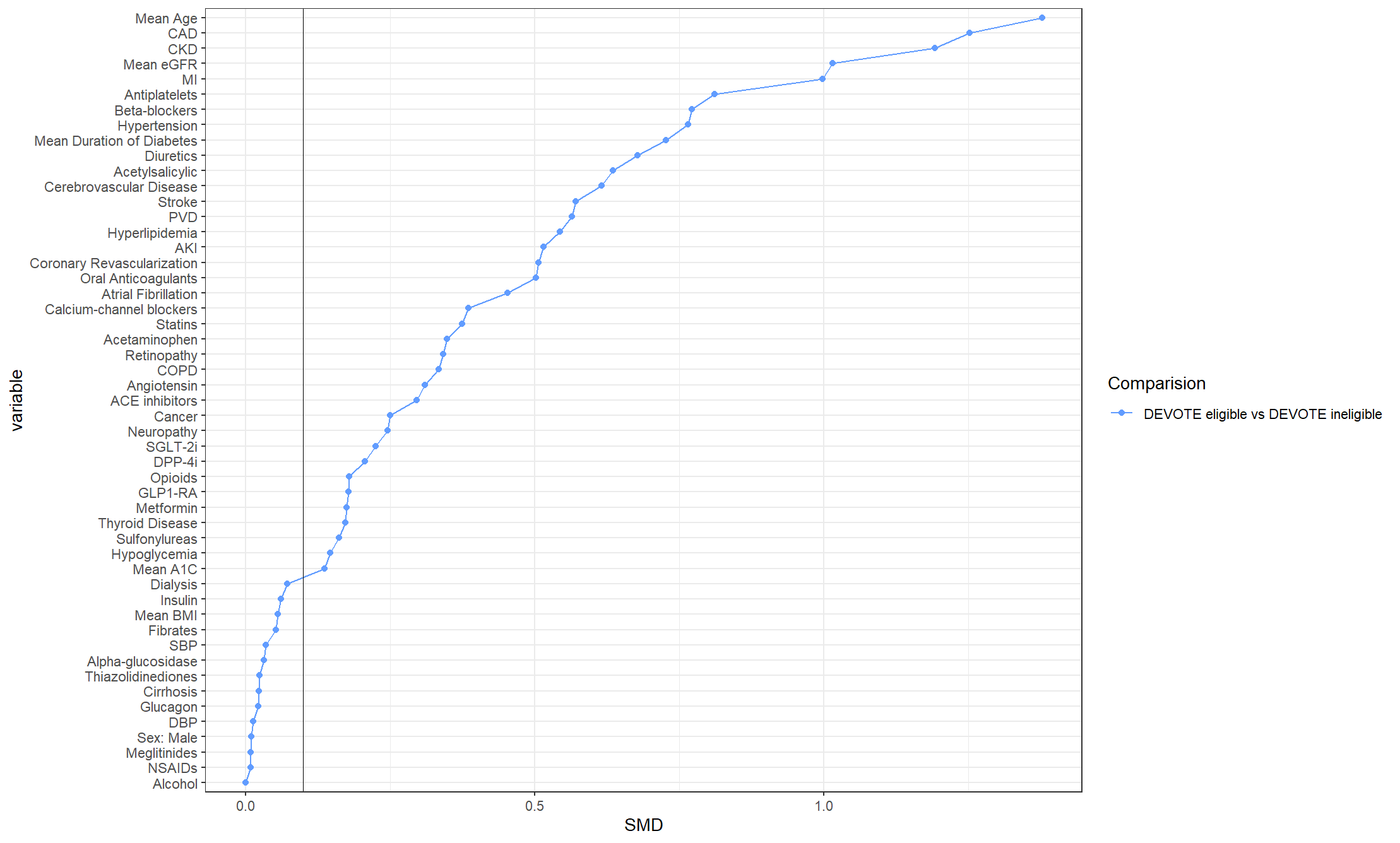
 Footnote: Graph depicts absolute standardized mean differences between DEVOTE eligible and DEVOTE ineligible total populations before weighting or imputation. Vertical line at 0.1 delineates clinically important differences

Abbreviations: A1C: glycated hemoglobin, ACE inhibitors: angiotensin-converting enzyme inhibitors, AKI: acute kidney injury, BMI: body mass index, CAD: coronary artery disease, CKD: chronic kidney disease, COPD: chronic obstructive pulmonary disease, DBP: diastolic blood pressure, DPP-4i: dipeptidyl peptidase-4 inhibitors, eGFR: estimated glomerular filtration rate, GLP-1 RA: Glucagon-like peptide-1 receptor agonists, IPTW: inverse probability of treatment weighting, MI: myocardial infarction, NSAIDs: Non-steroidal anti-inflammatory drugs, PVD: peripheral vascular disease, SBP: systolic blood pressure, SGLT-2i: sodium-glucose cotransporter-2 inhibitors, SMD: standardized mean difference

**Supplemental Figure S3** Plot of absolute standardized mean differences for patient characteristics in the DEVOTE trial compared to CPRD, DEVOTE eligible and DEVOTE ineligible prior to IPTW weighting and imputation

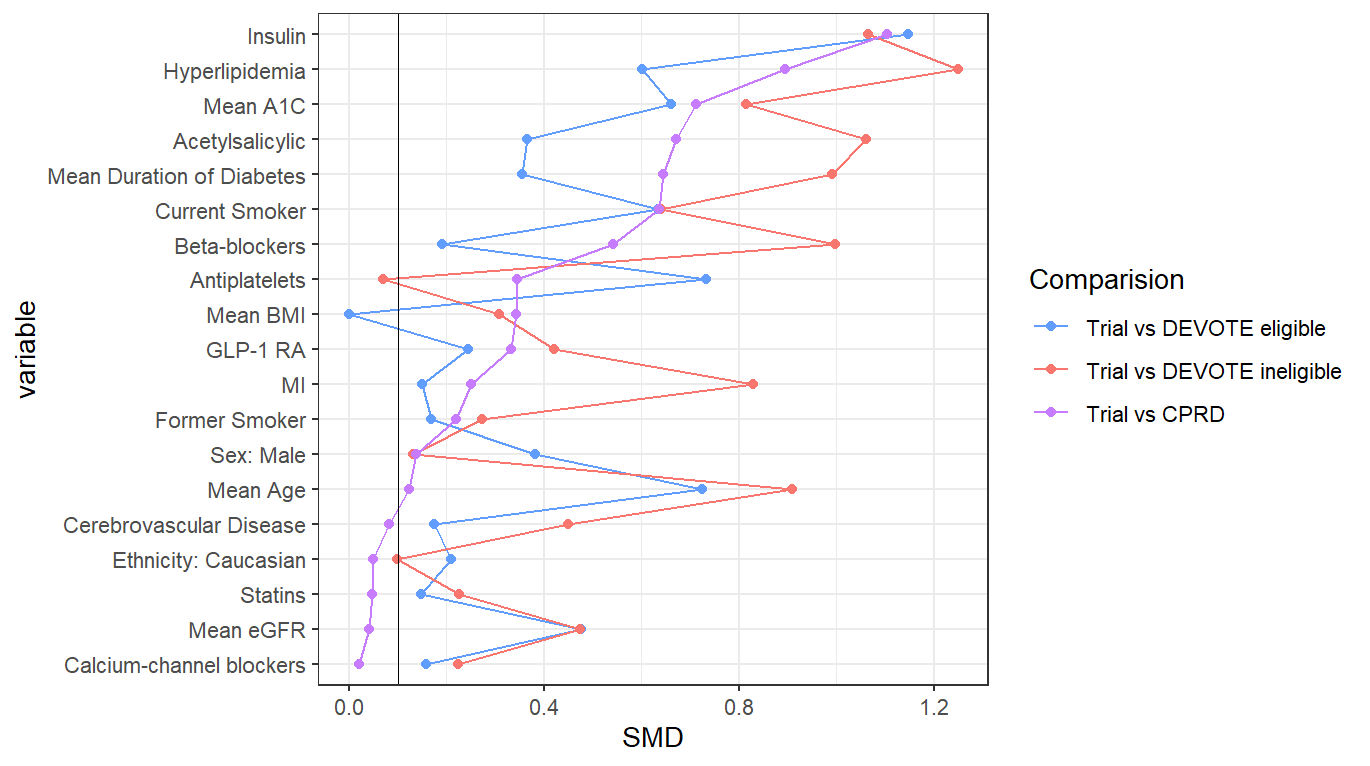

Footnote: Graph depicts absolute standardized mean differences between DEVOTE trial and CPRD, DEVOTE eligible and DEVOTE ineligible total populations before weighting or imputation in purple, blue and red respectively. Vertical line at 0.1 delineates clinically important differences.

Abbreviations: A1C: glycated hemoglobin, BMI: body mass index, CPRD: Clinical Practice Research Datalink, eGFR: estimated glomerular filtration rate, GLP-1 RA: Glucagon-like peptide-1 receptor agonists, IPTW: inverse probability of treatment weighting, MI: myocardial infarction, SMD: standardized mean differences
